# A mathematical model for tetanus transmission and vaccination

**DOI:** 10.64898/2026.03.16.26348506

**Authors:** Rachel A. Hounsell, Jared Norman, Sheetal P. Silal

## Abstract

Tetanus is a severe disease of the nervous system, transmitted through bacteria in the environment. In the absence of medical attention, case fatality rates are extremely high. Despite progress towards maternal and neonatal tetanus elimination targets, tetanus remains a serious public health problem. Routine infant and maternal vaccination have contributed to considerable reduction in cases and deaths from tetanus globally. However, protective immunity wanes over time. To increase duration of protection, the World Health Organization recommends three diphtheria-tetanus-pertussis-containing vaccine booster doses be given in early childhood, childhood, and adolescence. Evidence to support country-level decision-making about the introduction of these booster doses is critical. We have developed a novel age-structured, deterministic compartmental model of tetanus transmission and vaccination. The model is driven by environmental transmission and incorporates interventions like hygiene and safe birth practices to reduce the magnitude of environmental transmission. It explicitly models vaccination, separating each dose of the primary series, booster doses, and maternal vaccination to capture dose-specific effectiveness and duration of protection. The model captures heterogeneous immunity profiles by dose and age, and the cumulative nature of vaccine-derived protection. The immune dynamics follow the patterns described in literature and can replicate seroprevalence studies, although the exact characterisation of immunity in the literature still has gaps. This model presents a substantial advancement on previously published models and is well positioned to inform tailored vaccination strategies to reduce neonatal and non-neonatal tetanus.

## 1. Introduction

### 1.1. Biology and pathology of tetanus

Tetanus is a severe disease of the nervous system, characterised by muscle spasms and autonomic nervous system dysfunction [1]. Tetanus is not transmitted from person to person, but from bacteria in the environment [2]. While present in the environment throughout the world, the bacteria are most prevalent in hot and damp climates, where soil is rich in organic matter [3,4]. The bacteria typically enter the blood through contaminated wounds, releasing an extremely potent neurotoxin called tetanospasmin. Tetanus is typically characterised into neonatal and non-neonatal tetanus. Neonatal tetanus (NT), defined as tetanus in the first 28 days of life, is usually acquired at the site at which the umbilical cord is cut. Infection can occur due to poor sanitation protocols or cultural practices, such as the application of cow dung [5]. Despite the World Health Organisation (WHO) targeting maternal and neonatal tetanus elimination (MNTE) by 2000, it continues to be a public health concern [6].

Tetanus is diagnosed based on clinical features and does not require laboratory confirmation. Three clinical presentations characteristic of tetanus infection are: localised (uncommon), cephalic (rare), and generalised tetanus (most common, over 80% of cases) [3]. The overall severity of generalised tetanus disease and case fatality rates vary significantly depending on treatment, age, and general health of the patient [7]. In the absence of medical attention, case fatality rates (CFR) are extremely high [5]. A systematic review and meta-analysis of adult tetanus case fatality in Africa (27 studies from seven countries) estimated a pooled crude CFR of 45% [8]. A meta-analysis of 182 tetanus cases from 36 countries found a CFR in low- and middle-income countries (LMIC) of 30.3% (10/33) and in high-income countries of 19.4% (29/149), representing an odds ratio of 1.8 (CI:0.77- 4.19; p = 0.174) [9]. Management of tetanus cases includes 1) administration of human or equine tetanus immune globulin (TIG) to prevent further progression of disease; 2) antibiotics to prevent further progression; 3) benzodiazepines to control muscle spasms; and 4) supportive care to reduce risk of spasms [7]. Wound management for the prevention of tetanus includes passive immunisation using TIG and an age-appropriate tetanus toxoid-containing vaccine (TTCV) booster dose for those with incomplete or uncertain vaccination history. The distance from the wound site to the central nervous system is positively correlated with the incubation period and negatively correlated with disease severity [3]. The incubation period of non-NT usually varies between 3 and 21 days [3]. The incubation of NT is on average 7 days, with a wide interval [3,4].

### 1.2. Epidemiology of tetanus

A systematic analysis for the Global Burden of Disease (GBD) Study 2019 of the global burden of tetanus from 1990 to 2019 found significant decreases in tetanus incidence and death worldwide [10]. It estimated that there were 73,662 cases and 34,684 deaths from tetanus globally in 2019, down 88% from 615,728 estimated cases and 275,379 estimated deaths, respectively, in 1990 [10]. Despite these substantial decreases, tetanus is still a serious public health problem, especially in LMIC and countries with lower sociodemographic indices (SDI) [10]. A study on mortality from tetanus between 1990 and 2015 reported that 45% of NT deaths in 2015 occurred in South Asia, and 44% in Sub-Saharan Africa [11]. Of non-NT deaths, the authors estimated that 47% occurred in South Asia, 36% in sub-Saharan Africa, and 12% in Southeast Asia (Figure 1). The epidemiological profile of tetanus has evolved with the implementation of effective vaccination strategies. In countries with established immunisation programmes and high coverage rates, tetanus now predominantly affects older adults with waning immunity rather than neonates or children [10].

**Figure 1.**
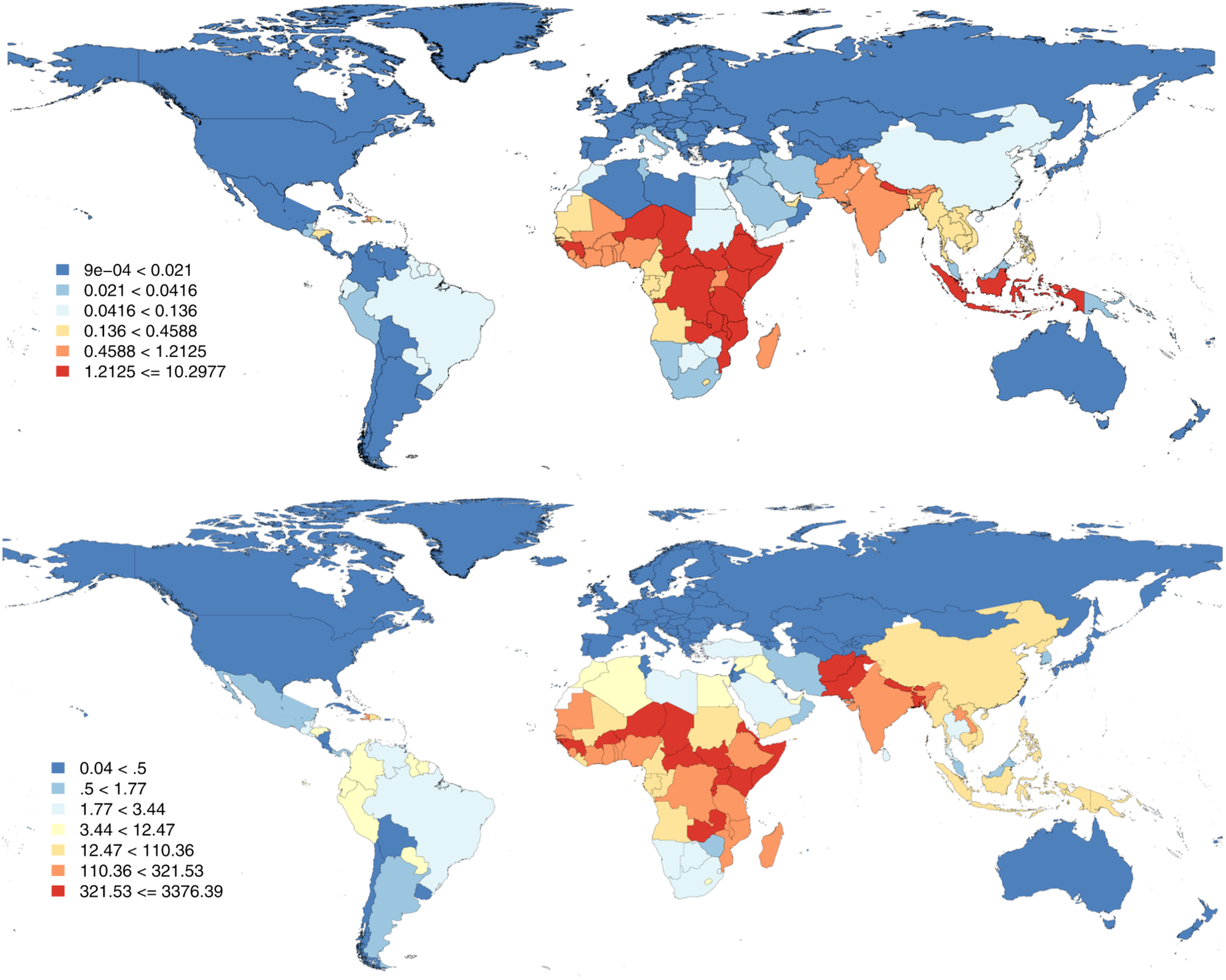
Neonatal tetanus (top map) and Non-neonatal tetanus (bottom map) mortality rate (per 100, 000 population), both sexes, 2015 [11]

The global neonatal tetanus elimination goal was launched at the World Health Assembly in 1989 to reduce neonatal tetanus as a public health problem (defined as less than one case of neonatal tetanus per 1000 live births in every district each year) in all countries [6]. The MNTE Initiative was launched by UNICEF, the WHO, and the United Nations Population Fund (UNFPA) in 1999, revitalising the goal and extending to including maternal tetanus elimination. This initiative employed a comprehensive strategy focusing on the immunisation of pregnant women and women of reproductive age with TTCV, promotion of clean delivery practices, and proper umbilical cord care. As of 2024, there were 10 countries that had not achieved MNTE [12]. However, some countries that had previously achieved MNTE have since reported ongoing NT cases and deaths. This emphasises the importance of continued political commitment, adequate funding, and innovative approaches to reach the most vulnerable populations to sustain these gains and achieve complete elimination.

### 1.3. Tetanus vaccination and immunity

There is no natural immunity to tetanus. Recovery from tetanus disease does not provide any protection against future infection. The only way to acquire immunity is through vaccination with tetanus toxoid (TT) [7]. TT induces the formation of specific antibodies called antitoxins, belonging to the immunoglobulin G (IgG) class, which circulate in the bloodstream and extravascular spaces [2]. These antitoxins can neutralise the toxin produced in a wound infected with *C. tetani* and, when passed to a foetus through the placenta of a vaccinated mother, can prevent NT. Establishing a definitive threshold for the protective level of tetanus antibodies has been subject to much debate in the literature. Some studies use a minimum level of antibody required for protection of 0.01 UI/ml, measured by an in vivo neutralisation assay. However, others use a higher cutoff of 0.1 IU/ml. Even at these thresholds, several studies have reported cases of tetanus occurring in persons with antibody levels greater than or equal to 0.01 IU/mL by neutralisation assay, or 0.16 IU/mL by ELISA [2]. This illustrates the difficulty in determining a protective antibody level, further exacerbated by the use of different techniques to measure antibody response. While usually well correlated, there can be large differences in sensitivity, specificity, and accuracy across techniques. As such, guidelines stipulate that the test method, cutoff used, and the correlation with a standard validation process should be reported alongside the tetanus IgG assay results [2].

Development of immunity following vaccination depends on the age when given and the timing of doses relative to one another [2]. A single dose of TT is insufficient to induce protective levels of antibodies. During the primary series of DTPCV for infants, the protective threshold for tetanus is usually exceeded 2–4 weeks after the second dose. The third dose induces protective immunity in almost 100% of those vaccinated. Immunity wanes over time, with each additional dose extending the duration of immunity. Figure 2 provides a schematic diagram of the typical immune response following one to five TTCV doses. Data from serological studies suggest that the full primary series plus a booster during the second year of life will provide 3–5 years of protection [7]. Others suggest that the fourth dose (given in the second year of life) may extend protection up to ten years [2]. The childhood booster dose, offered at 4–7 years of age, is estimated to provide protection for 10–20 years, while the adolescent booster dose, offered at 9–15 years of age, is likely to provide immunity that lasts through most of adulthood (20–30 years or longer). Vaccination with at least two doses of TT during pregnancy is estimated to reduce mortality in neonatal tetanus by 94% [4], and provide protection for the mother for 3–5 years [7]. The exact level, duration and pattern of waning of immunity is not well defined.

**Figure 2.**
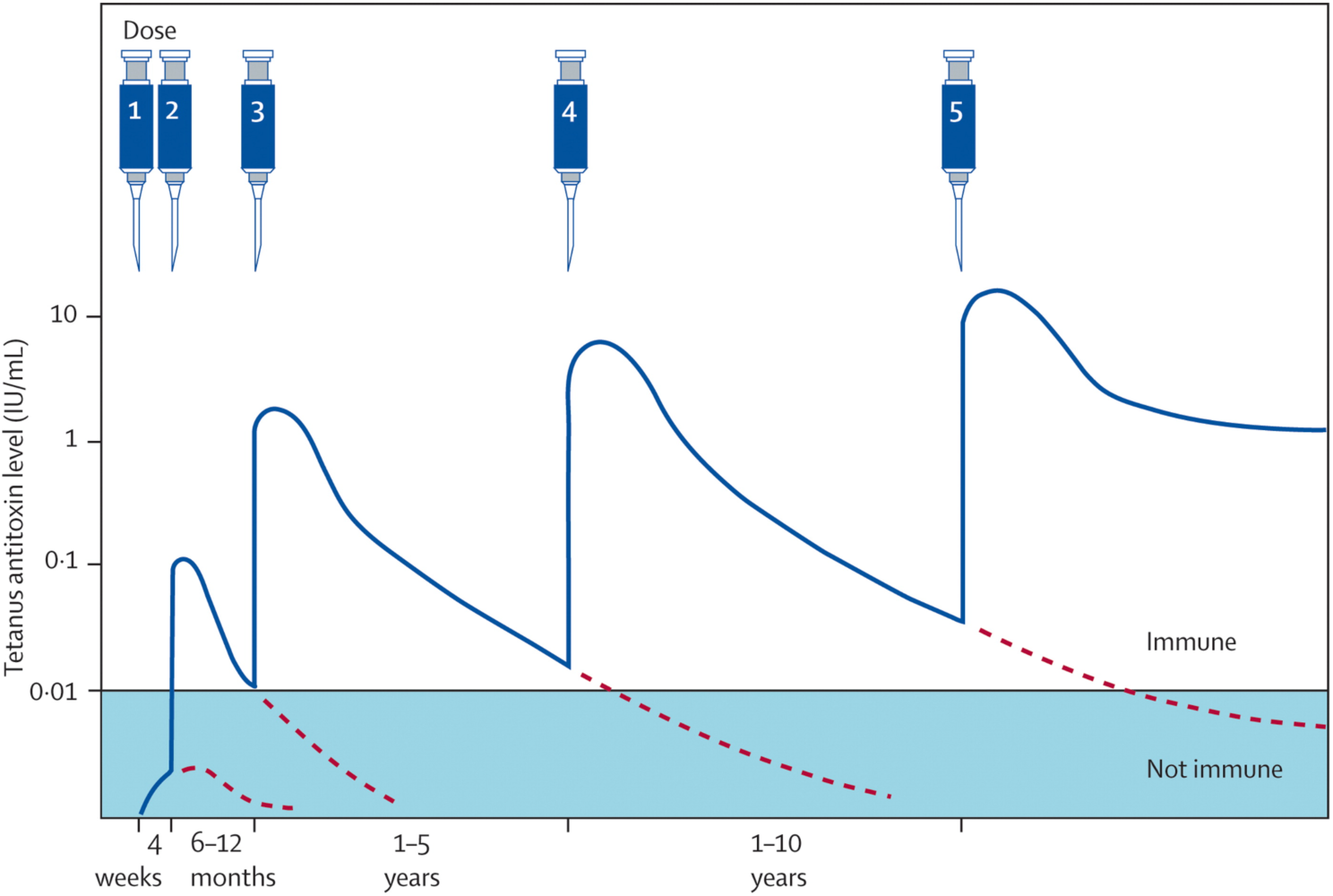
Antibody response to tetanus toxoid [2,5,13]

### 1.4. Mathematical models of tetanus

There are very few epidemiological models of tetanus in published literature. The most comprehensive model is Cvjetanovic et al. (1972 and 1978) [14,15]. This model is a dual SEIRD (susceptible-exposed-infectious-recovered-dead) model accounting for both NT and non-NT in the general population. The aim of the model was to support the planning of immunisation programmes and control strategies. The force of infection in this model accounts for the prevalence of *C. tetani* in the environment, the frequency of lesions, the proportion of the population that is rural, socioeconomic conditions, and the level of medical care for the wounded and during deliveries. Another model, by Carducci et al. (1989), estimates the epidemiological and economic effects of different tetanus vaccination strategies [16]. The authors conduct a cost-benefit analysis of increasing vaccine coverage in over 10-year-olds in Tuscany. The model considers the population in age bands of 10 years, starting at 3 months of age when the first vaccination starts. The authors elect not to model neonatal tetanus, as it is exceedingly rare in Italy.

Carter et al. (2024) developed a statistical model for tetanus in their work to estimate deaths averted due to vaccination against 14 pathogens in 194 countries from 2021 to 2030 [17]. For tetanus, they used a logistic regression model to extrapolate the relative risk of death under different vaccination assumptions. While an appropriate approach for their analysis, the model is not suitable for exploring population immunity over time, or able to project cases and deaths by age group. Further, it only estimates the impact of the primary series and does not account for potential impact of DTPCV booster doses or maternal vaccination. This model was extended by Shattock et al. (2024) to include a booster schedule and maternal vaccination in their analysis to estimate the contribution of vaccination to globally declining infant and child mortality rates since the introduction of the EPI in 1974 [18].

Several studies have conducted economic evaluations of DTPCV [19–24]. However, most either focused solely on the impact related to pertussis [19–23] or—where a tetanus component was included—conducted simple assumption-based analyses to estimate burden changes using a linear vaccine effectiveness multiplier [24]. Two notable economic models for tetanus vaccination exist. Griffiths et al. (2004) used a state-transition model to estimate the incremental cost-effectiveness of supplementary immunisation activities to prevent neonatal tetanus in Pakistan [25]. Laing et al. (2020) used a statistical model to inform an investment case for MNTE in 13 countries yet to achieve elimination [26]. This paper contributes to the literature on tetanus modelling by developing an age-structured, deterministic compartmental modelling capturing population immunity, tetanus incidence and deaths (both NT and non-NT), with vaccination by dose (primary series, booster doses, and maternal vaccination).

## 2. Materials and methods

This section presents an overview of the age-structured, compartmental tetanus model, key assumptions, and parameters. Model development and parameters were informed by engagement with a technical expert group (TEG). Figure 3 shows the conceptual model diagram for tetanus with a list of corresponding model compartments. Table 1 and Table A.3 (Appendix) show the model parameters for tetanus.

**Figure 3.**
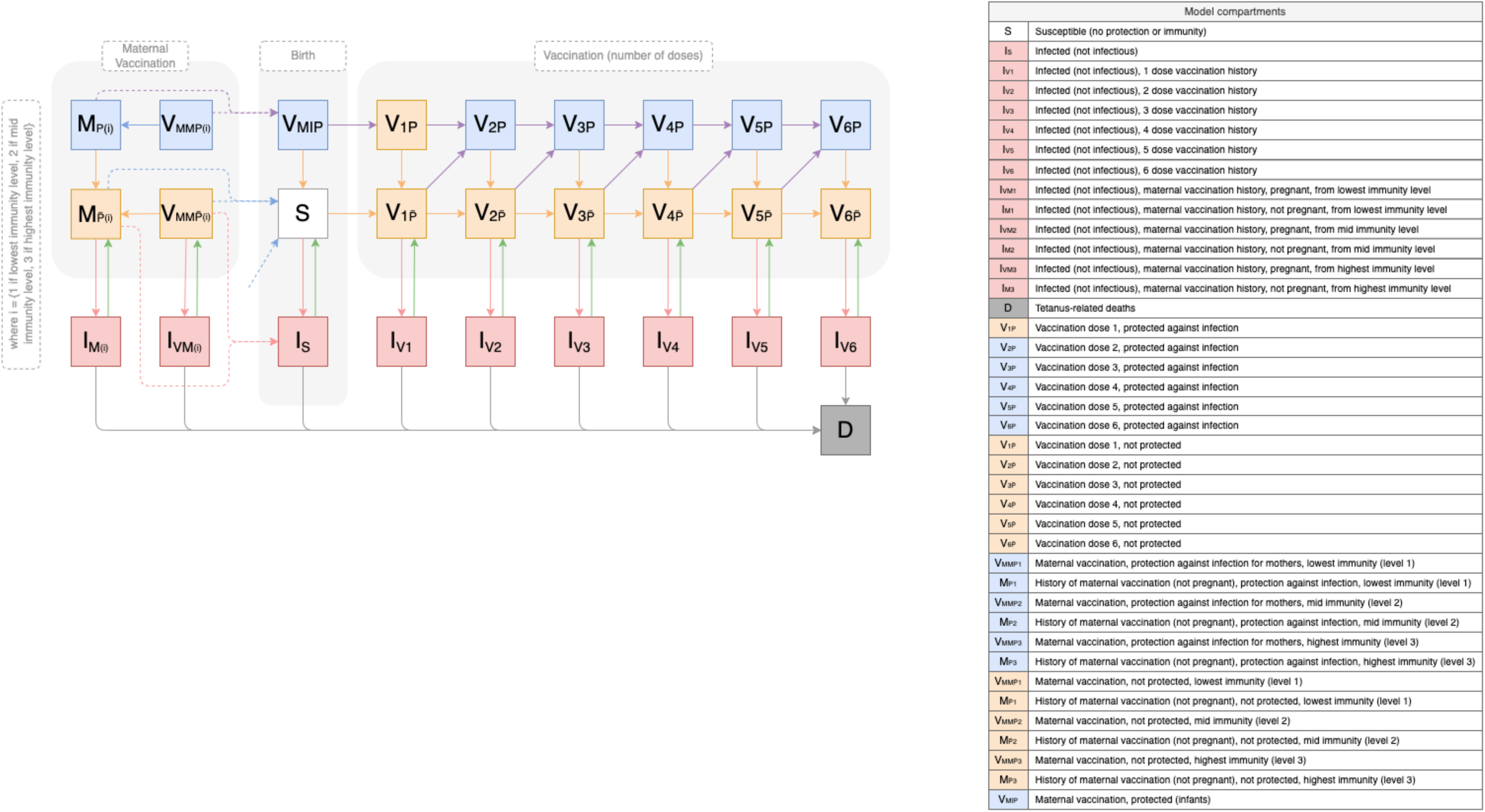
Conceptual diagram for the tetanus model structure

**Table 1.** Description and dimensions of model parameters for tetanus.

| Parameter | Symbol | Description | Time-dependent | Age specific | Infection type specific |
| --- | --- | --- | --- | --- | --- |
| Demographic parameters |  |  |  |  |  |
| Fertility | $births(t)$ | Number of births | Y | Y | N |
| Aging | $\alpha_i$ | Age rate: Progression through the 56 model age stratifications $i$ | N | Y | N |
| Mortality | $\mu_i(t)$ | All-cause mortality | Y | Y | N |
| Contact matrix<sup>^</sup> | $C$ | An $na \times na$ matrix where the $(i,g)^{th}$ entry is the number of contacts that an individual in Age Group $i$ has with individuals in Age Group $g$ (in chosen units) | N | Y | N |
| Biological and clinical pertussis parameters |  |  |  |  |  |
| Mortality (tetanus-related) | $\mu_{T_t}$<br>$\mu_{T_{nt}}$ | Tetanus mortality: Percentage of treated (**t**) or untreated (**nt**) cases resulting in death | N | N | N |
| Recovery | $\gamma$ | Recovery rate from infectious states to R: 1/(average duration of infectiousness) | N | N | N |
| Transmission scalar | $\phi$ | Calibration parameter for the force of infection in the environment | N | N | N |
| Vaccine-related parameters |  |  |  |  |  |
| Waning of vaccine-induced immunity ($V_P$ to $V_{NP}$) | $\tau_v$<br>$\forall v \in \{1,2,3,4,5,6\}$ | Waning rate from $V_P$ to $V_{NP}$ (1/average duration of protection) for vaccine states $V_{1P}, V_{2P}, V_{3P}, V_{4P}, V_{5P}, V_{6P}$ | N | N | N |
| Waning of immunity from maternal antibodies | $\tau_{mm}$<br>$\tau_{mi}$ | Waning rate from $M_{x,P}$ to $M_{x,NP}$ (1/average duration of protection of maternal antibodies in mothers) for prior immunity level $x = 1$ (lowest) to 3 (highest) | N | N | N |
| | | Waning rate from $V_{MI,P}$ to $S$ (1/average duration of protection of maternal antibodies in infants) | | | |
| Vaccine effectiveness | $\epsilon_v$<br>$\forall v \in \{1,2,3,4,5,6\}$ | Vaccine effectiveness (population fully protected against infection) for each corresponding dose | N | N | N |
| Vaccine effectiveness for maternal vaccination | $\epsilon_{M1-3}$ | Vaccine effectiveness (proportion of infants fully protected at birth through maternal vaccination antibodies) for prior immunity levels 1 (lowest) to 3 (highest) | N | N | N |
| DTPCV intervention parameters |  |  |  |  |  |
| Age-specific vaccine coverage | $cov_{v,l}(t)$<br>$\forall v \in \{1,2,3, b, cb, ab\}$ | Derived based on inputs:<br>Ages when eligible to receive vaccine<br>Proportion who successfully receive vaccine at each eligible age ($cov_{v,i}$ equals 0 for Age Group $i$ if there is no vaccination at age $i$) for each year | Y | Y | N |
| Maternal vaccine coverage | $cov_m(t)$ | Proportion of pregnant women successfully vaccinated (intervention scenario input) | Y | Y | N |

Notes: ^The contact rate does not affect transmission in the model. Abbreviations: Y, yes; N, no; DTPCV, diphtheria, tetanus, pertussis-containing vaccine.

### 2.1. Model structure

The model is a compartmental transmission model with 36 states, stratified by 56 age groups (*i*). Age categories were chosen to simulate vaccination and immunity at key ages; from very fine age groups (several weeks or months during the first two years of life), to annual age groups through to 15-years old, then five-year age bands until >75-years old. For simplicity, age subscripts are not included in the model description below but are shown in the model equations for clarity. The full list of model age groups (Table A.1), model structure details by compartment (Table A.2), and model equations are provided in the Appendix.

As tetanus is not transmitted from person to person, but from bacteria in the environment, those in the infected state (I_j_) are infected only, not infectious. The force of infection for non-neonatal tetanus is driven by a transmission scalar, which is used to calibrate the model to the annual number of tetanus cases in the chosen setting. Where reported cases are used for calibration, these are first adjusted by a reporting factor. As clinical presentation (localised, cephalic, and generalised tetanus), duration and severity of disease do not have an influence on immunity or onwards transmission, these are not specified in the compartmental model. There is a single type of infected state (I_j_), which is differentiated based on the model state *j* prior to infection. This maintains vaccination history in the model, such that the vaccination schedule may continue after infection. Given that infection does not lead to any naturally acquired immunity, the infected population recovers back to the model state they were in prior to infection. Those who die from tetanus move into D (disease-related death compartment) at the disease-related mortality rate (μ_T_). Individuals exit the model at an age-specific natural mortality rate (μ_i_).

Immunity to tetanus is only acquired through vaccination with a TTCV dose. Therefore, vaccination drives all tetanus immunity in the model. Vaccination protects against infection, not just disease. The vaccinated states in the model cover the six WHO-recommended DTPCV doses (three primary series and three booster doses) and maternal immunisation. As vaccine effectiveness and duration of immunity for tetanus is dependent on the cumulative number of TTCV doses, the model states are captured as six sequential, cumulative doses (V_1_-V_6_). Therefore, the eligible population would only receive higher doses in the schedule if they have received the previous doses (e.g. can only move into V_2_ if they move from V_1_, not S). The first received TTCV dose provides no protection against tetanus. Therefore, irrespective of which DTPCV dose is first (e.g. if an infant does not receive the primary series but goes on to receive a booster dose later in childhood), the model will capture this as V_1_, with no protection offered against tetanus infection. For each subsequent dose received, the eligible population will move into the next vaccine dose state.

Vaccination occurs on ageing where the age-stratified population either move into the next age group in the same state if no vaccine is received, or into the next age group of the appropriate vaccinated state, depending on the dose-specific coverage in the given year. The age at which each vaccine is offered depends on the national schedule. The 56 age groups allow for the variation in schedules between countries to be captured and scenarios simulating different ages for the introduction of each booster dose. For each vaccine dose, we assume that those moving from a protected vaccination state to a higher dose state will remain protected by the vaccination (e.g. V_2P_ will only ever move into V_3P_, not V_3<u>P</u>_), as one should not be able to become less protected after receiving another dose. However, those moving from a non-protected state (either from S or from an unprotected vaccinated state) may move into the protected or non-protected state of the subsequent dose. This is based on the corresponding dose’s effectiveness (ε) (e.g. proportion ε move from V_2<u>P</u>_ to V_3P_ and proportion (1 – ε) move from V_2<u>P</u>_ to V_3<u>P</u>_). Over time, vaccine-derived immunity wanes at a dose-specific rate, allowing individuals in the protected vaccinated states to transition to the corresponding non-protected vaccinated state (e.g. V_2P_ to V_2<u>P</u>_). From the non-protected vaccinated states, individuals are fully susceptible to infection.

Maternal vaccination is modelled to be offered to pregnant women based on age-specific fertility rates via antenatal care visits as one TTCV dose (TTCV1) or with two or more doses (TTCV2+). Protection following maternal vaccination is modelled through twelve states for mothers and one state for infants, who are protected by maternal antibodies after birth. The twelve states for mothers are stratified into three levels of protection: lowest immunity (level 1), mid immunity (level 2) and highest immunity (level 3); and four states capturing whether the mother has been vaccinated for the current pregnancy or a previous pregnancy, and whether this provided protection, based on vaccine effectiveness. The immunity level (1 to 3) following maternal vaccination then depends on the prior state of immunity (history of vaccination with the primary series and booster doses) and the number of TTCV doses received for maternal vaccination. This allows the model to retain the level of immunity and corresponding duration of protection. Individuals move from V_MMP*k*_ and V_MM<u>P</u>*k*_ into M_P*k*_ and M<u>_P_</u>*_k_* respectively after the nine-month period of pregnancy. This captures the history of maternal vaccination and allows women to retain vaccine-derived protection beyond the period of pregnancy. Individuals move from a protected state (M_P*k*_) into an unprotected state (M<u>_P_</u>*_k_*) based on the waning rate of the corresponding immunity level.

Individuals are born into three potential states in the model. Infants who are born to mothers with immunity from a protected vaccinated state are assumed to possess protective antibodies at birth (V_MIP_). Infants born to mothers without immunity are either infected with neonatal tetanus, depending on the neonatal tetanus transmission scalar, upon birth (I_s_) or are born into a fully susceptible state (S).

### 2.2. Model parameters

Table 1 shows the description and dimensions (time, age, and infection type) of the model parameters for tetanus. Table A.3 provides the unit, range, distribution, and source of the parameter values for tetanus. Parameter values were informed by literature and discussions with disease experts on the TEG.

### 2.3. Sensitivity analysis

Drawing from the appropriate parameter distributions (Table A.3), we conducted Latin hypercube sampling (LHS) of the parameter space to generate 1000 parameter sets. As the immunity waning parameters have overlapping ranges, we combined them into three groups of waning parameters for the purpose of LHS: immunity waning rate of doses 1–3, dose 4, and doses 5–6. LHS for the immunity waning parameters was drawn from a uniform distribution between one and three for each set, where two represents the default value, one the minimum duration of protection, and three the maximum duration of protection. This approach enabled us to distinguish between the primary series and booster doses for the sensitivity analysis, without resulting in parameter sets that contravened the principle of increasing immunity following each subsequent dose.

After running the model 1000 times on each of the LHS parameter sets and obtaining the corresponding model outputs, we calculated the partial rank correlation coefficient (PRCC) using the epiR package (version 2.0.80) in R. Input variables for the PRCC are the tetanus parameters varied in the LHS: Tetanus transmission scalar, likelihood of death for an untreated case, likelihood of death for a treated case, the tetanus recovery rate, vaccine effectiveness, immunity waning rate (Doses 1–3), immunity waning rate (Dose 4), and immunity waning rate (Doses 5–6). Three model outputs (average annual tetanus incidence, proportion of the population protected against tetanus, and average annual number of tetanus deaths) for three age groups (all ages, under 5-year-olds, and under 15-year-olds) are considered for the PRCC.

## 3. Results

### 3.1. Sensitivity analysis findings

For tetanus incidence, there are three input parameters with a significant PRCC: tetanus transmission scalar, vaccine effectiveness, and immunity waning rate (doses 1–3). The transmission scalar is the highest ranked, followed closely by the immunity waning rate (doses 1–3). This is consistent across the three age groups considered. Figure 2.6 shows the interaction between these two highest ranked variables in relation to tetanus incidence. As the transmission scalar is used to calibrate to the chosen setting, the exact range of values is determined by the transmission potential in the environment and the number of reported or estimated cases (i.e. the transmission scalar does not have a predetermined ranged based on literature). A subset of values would be selected based on model fitting (an example is illustrated by the grey shaded band in Figure 4).

**Figure 4.**
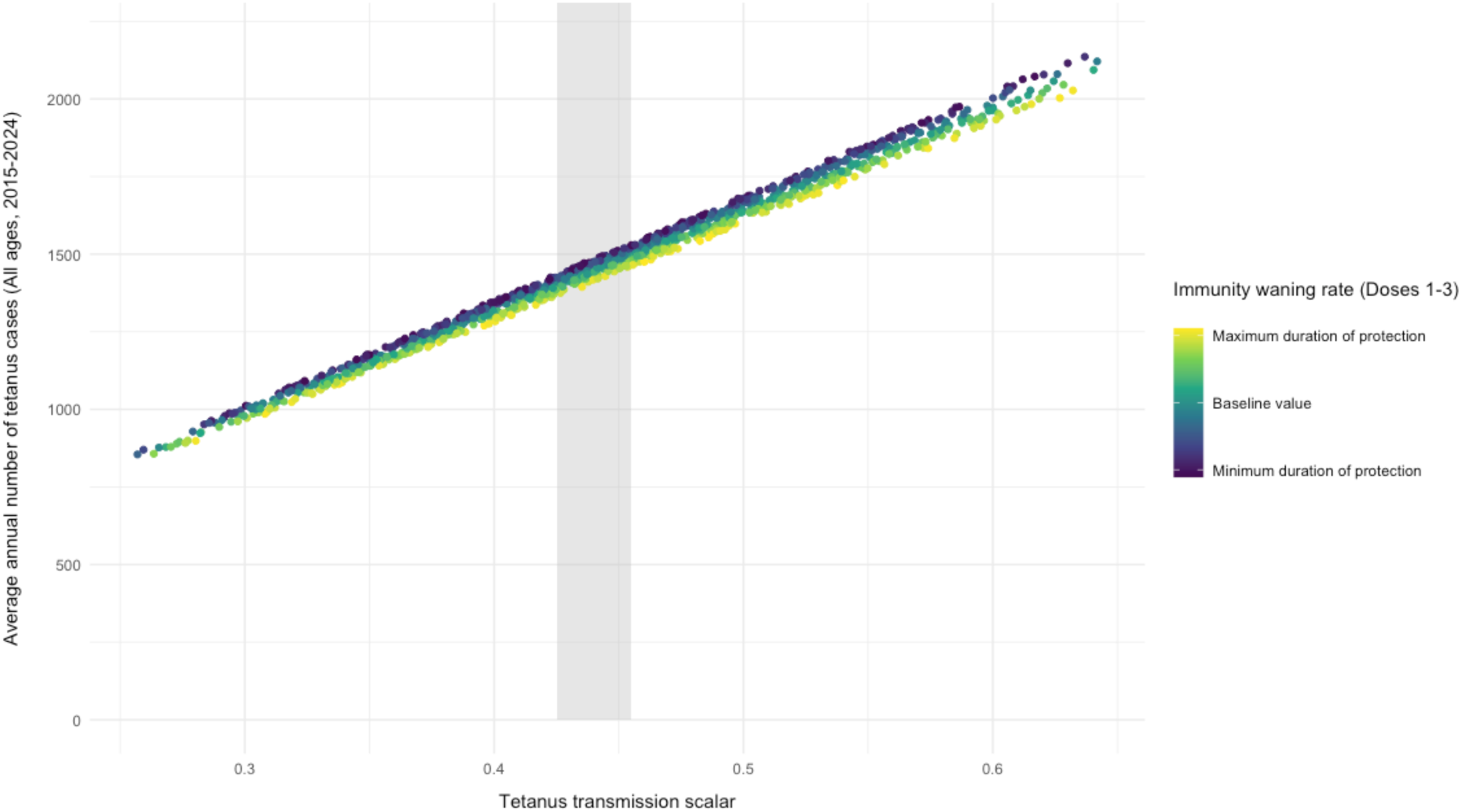
Projected average annual number of tetanus cases in all ages for each of the 1000 LHS parameter sets, shown in relation to the tetanus transmission scalar and immunity waning rate for TTCV doses 1–3 Figure description: Each point represents a model run with one of the 1000 LHS sets. The points are coloured according to the range of values for the immunity waning rate of TTCV doses 1–3 (from dark purple representing the minimum modelled value to yellow representing the maximum modelled value for duration of protection against tetanus infection). See Table A.3 for ranges and sources. Abbreviations: LHS, Latin hypercube sampling; TTCV, tetanus-toxoid-containing vaccine.

Figure 5 shows the PRCC from 1000 model runs of a scenario with the primary series only (no booster doses). The bars are coloured to make the input variables easily distinguishable across the nine panels. The black lines show the 95% confidence interval (CI) for the PRCC. Where the CI crosses zero, the PRCC for the corresponding input parameter is not statistically significant at the 5% level.

**Figure 5.**
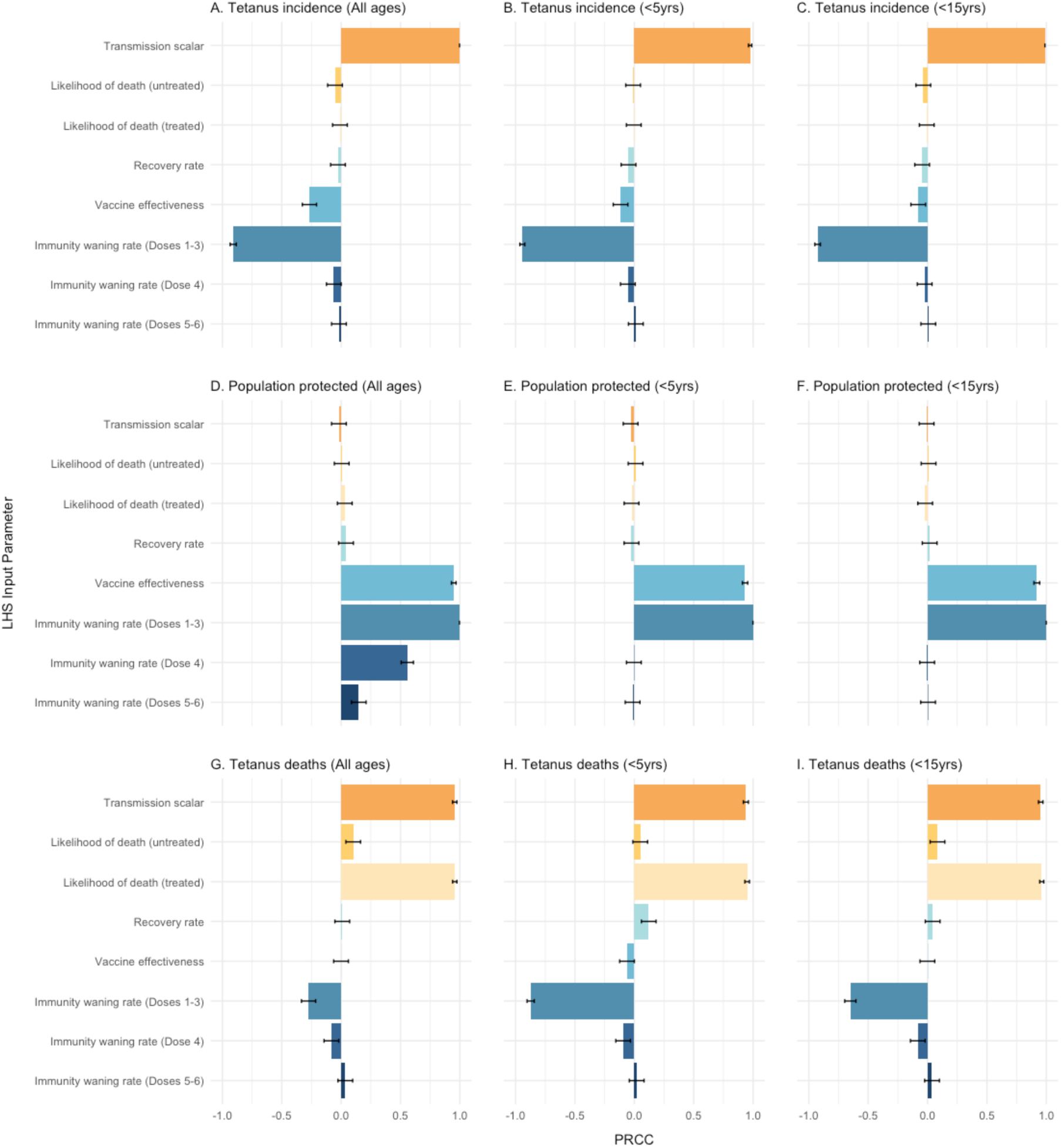
Partial rank correlation coefficient (PRCC) with 95% confidence interval of key tetanus parameters in relation to tetanus incidence, population protected and deaths, for the DTP primary series (1000 model runs) Figure description: PRCC of tetanus parameters in relation to tetanus incidence (Panels A – C), proportion of the population protected (%) (Panels D – F), and tetanus deaths (Panels G – H). The sensitivity analysis is subset by age: All ages (Panels A, D, G), children under 5-years of age (Panels B, E, H), and children under 15 years of age (Panels C, F, I). PRCC values above zero are positively correlated with the output variable and those below zero are negatively correlated with the output variable. The 95% confidence interval is shown in the black whiskers on each bar. The values are drawn from 1000 model runs. Abbreviations: LHS, Latin hypercube sampling; PRCC, partial rank correlation coefficient.

For population protected, immunity waning rate (doses 1–3) and vaccine effectiveness (first and second highest ranked) are significant with the highest ranking of the PRCCs across all three age groups. Additionally, for the analysis on all ages, the immunity waning rates for dose 4 and doses 5–6 are also significant, although lower ranked (third and fourth largest effect) than for doses 1–3. This makes sense because, in a scenario with the primary series and maternal vaccination only, the population under fifteen years of age will have only been offered DTPCV doses 1–3 (primary series), while across the whole population, doses 4 to 6 are offered through maternal vaccination.

When estimating tetanus deaths, the model is most sensitive to the transmission scalar and the likelihood of death. The third highest ranking for tetanus deaths is the immunity waning rate for doses 1–3, which is negatively correlated with deaths, most prominently in the population under five years of age.

### 3.2. Immunity dynamics

There is still much uncertainty in the literature about the exact nature and duration of protection that TTCV vaccines provide against tetanus. For compartmental models, a standard approach to incorporating waning rates is the use of exponential decay. However, as the duration of immunity is one of the most sensitive model parameters (Figure 2.6), it is important to understand the impact of different approaches on how this is represented in the model. To do this, we modelled two approaches (threshold versus exponential), using a hypothetical setting with the primary series and an assumed duration of protection for three doses of five years. Figure 6 shows the comparison of the projected proportion of the population protected against tetanus in the model using these two different approaches for modelling immune waning. The threshold approach assumes that protection is binary (i.e. individuals are protected above the 0.1 IU/mL cutoff and not protected once they fall below this level). In this case, the cohort receiving the primary series (after accounting for vaccine effectiveness) will be fully protected for five years, after which they lose all protection and transition to the corresponding not-protected vaccine compartment. For the exponential approach, the population starts at the same proportion of protection and for each year since vaccination, this proportion declines exponentially.

**Figure 6.**
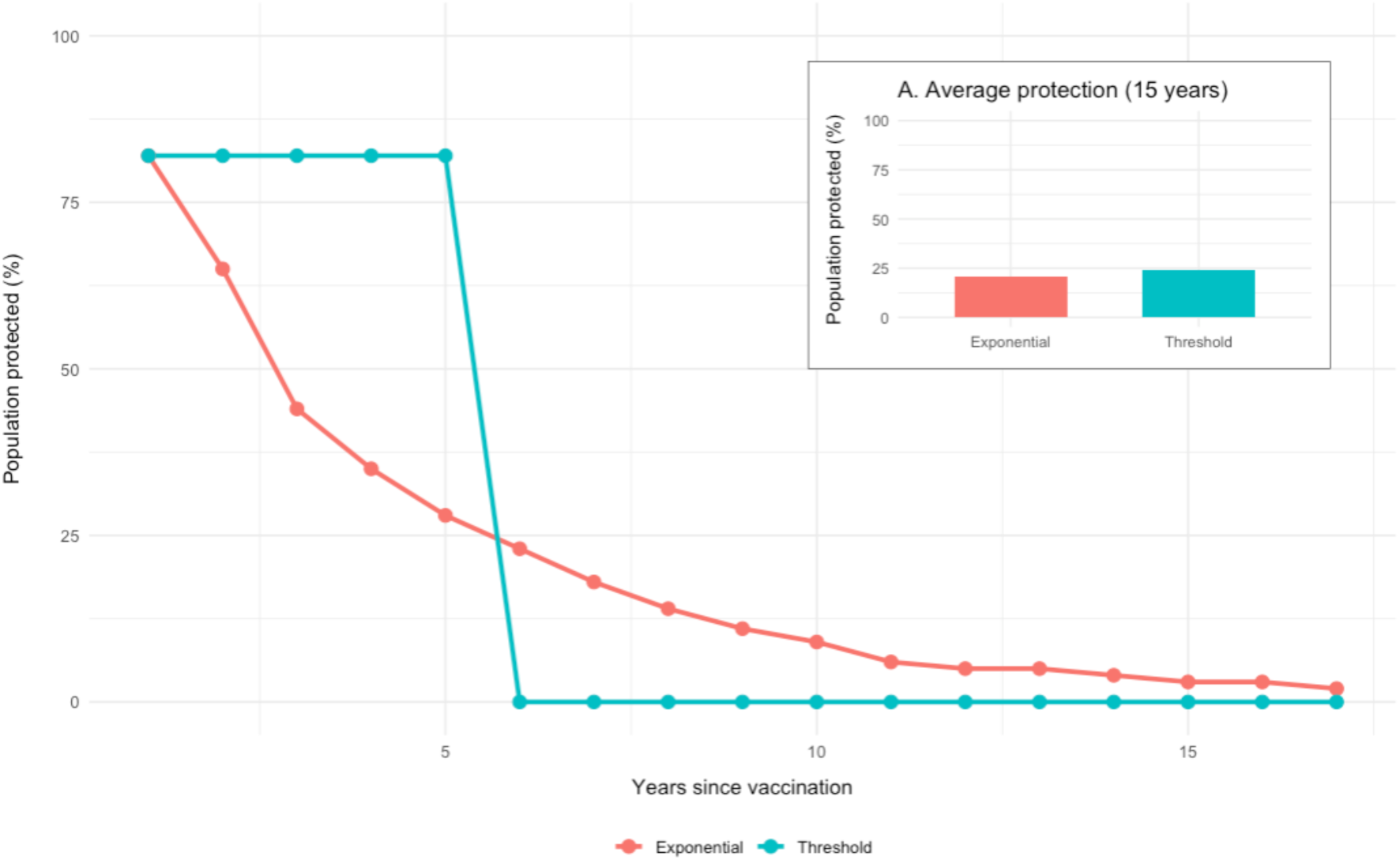
Comparison of two approaches for modelling the waning of tetanus immunity following primary series vaccination

The model outputs show that, although the pattern of decline is different, the two approaches are comparable in terms of average proportion of the population protected after 15 years following the last dose—21% for the exponential approach versus 24% for the threshold approach. The results will also be comparable in the overall population (all ages). However, using the exponential approach, results for the proportion protected in the under 5-year-old population will be lower than the threshold approach.

A third approach, which we did not model, would be the application of a waning function (e.g. a sigmoid curve) that is somewhere in between, where there is full protection in the first five years, after which there is a decline. In a setting where there is local seroprevalence data by age, the function can be fitted to represent the observed decline in immunity. In the absence of this data, we use the exponential waning approach and present results in the under 15-year-olds and all ages.

Figure 7 shows the projected proportion of the population that have protective immunity against tetanus following the primary series, in annual age bands from 0 to 15 years of age, as well as summary age groups for the under 5-year-olds, under 15-year-olds, and all ages. Given uncertainty in the literature regarding the duration of immunity following each DTPCV booster dose, the proportions are shown as boxplots, capturing the range in values drawn from the 1000 model runs. Figure 8 shows the same output under scenarios of one (EC), two (EC+C and EC+A), and three (EC+C+A) DTPCV booster doses.

**Figure 7.**
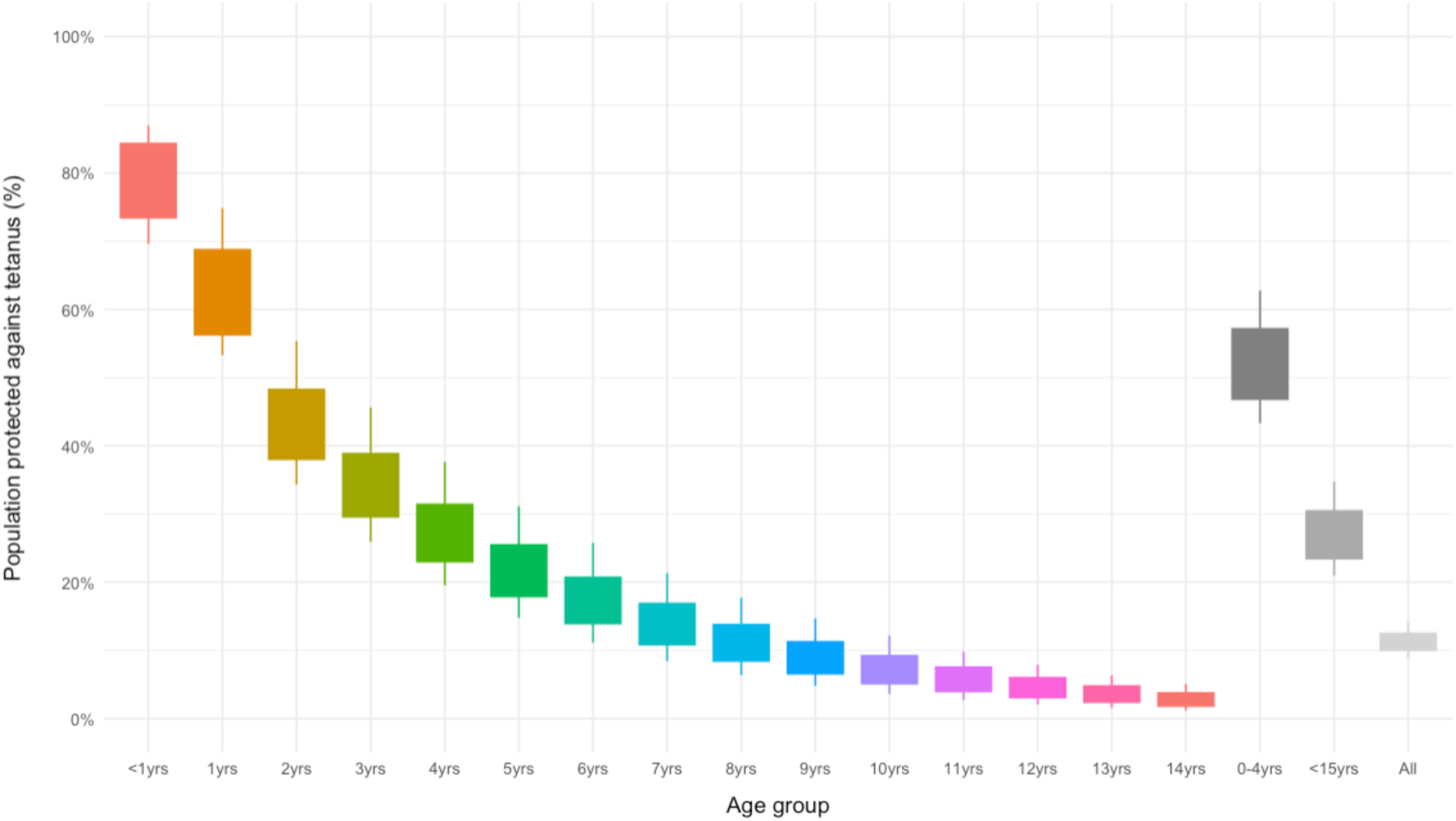
Projected proportion of the population protected against tetanus, by age group, following vaccination with the primary series only

**Figure 8.**
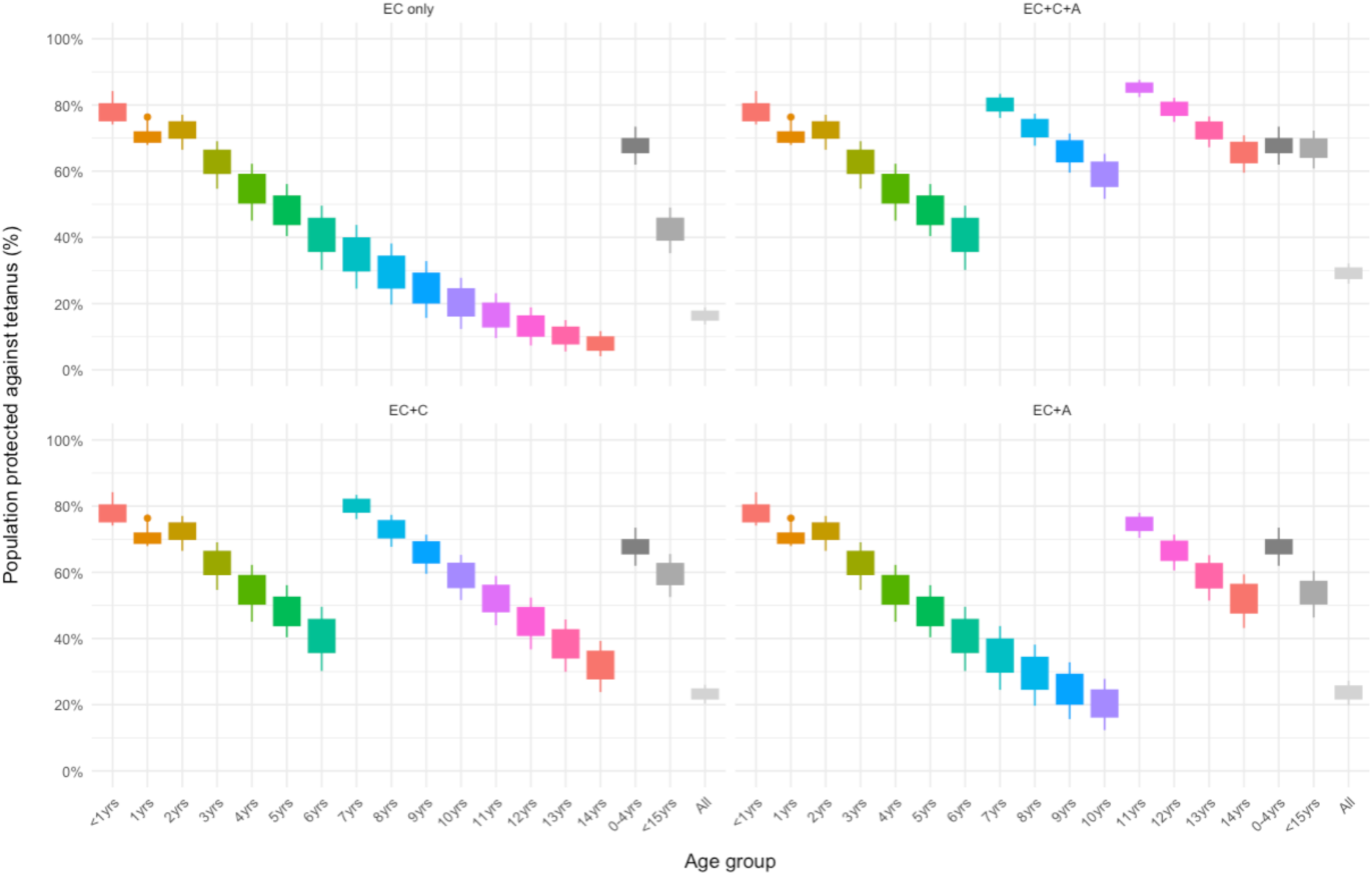
Projected proportion of the population protected against tetanus, by age group, for different combinations of DTPCV booster doses

The pattern of immune waning qualitatively aligns with those described in literature [2,7], as shown in Figure 2. That is, it produces the expected pattern of decline following vaccination. However, to validate quantitatively that the model reproduces realistic ranges of population protection, we compared model outputs with seroprevalence studies on tetanus (Table 2). We selected three studies that cover a range of age groups and settings in terms of number of doses and coverage levels. The first is Tohme et al. (2023), who studied tetanus and diphtheria seroprotection among children younger than 15-years in Nigeria—a setting with only the primary series [27]. The second is Liu et al. (2021), who assessed the serologic immunity to diphtheria, tetanus and pertussis in China—a setting with four TTCV doses—in the population up to 89 years of age [28]. The third is Pachon et al. (2002), who assessed the age-specific seroprevalence of poliomyelitis, diphtheria and tetanus antibodies in Spain—a setting with five TTCV doses—in the population 2 to 39 years of age [29]. For the validation, we ran the tetanus model with the vaccine schedule and coverage from each study setting. We then calculated the proportion of the population protected against tetanus projected by the model for the relevant age categories. Although the age categories from the seroprevalence studies do not map exactly onto those in the model, we estimated the equivalent age categories by assuming equal distribution of immunity within the age bands. For all studies, we have used the protective cutoff of 0.1 IU/mL to compare against the model outputs. We present the estimate as well as the range for the upper and lower bounds of the duration of protection for the relevant vaccine doses. All other parameters remained constant.

**Table 2.** Validation of modelled population protection.

| Study setting and description | Age groups | Study seroprotection (%)<br>>0.1 UI/mL | Model population protection (%) |
| --- | --- | --- | --- |
| 3 Doses (Primary series) |  |  |  |
| Tohme et al. (2023) [27]<br>Setting: Nigeria<br>Ages included: 0–15 years<br>Cross-sectional study (n = 31,456) | <15y<sup>^</sup> | 26.4% [21.4, 31.9] | 21.9% [15.2, 30.4] |
| 4 Doses (Primary series + early childhood booster) |  |  |  |
| Liu et al. (2021) [28]<br>Setting: China<br>Ages included: 0–89 years<br>Cross-sectional study (n = 666) | 3m – <18m | 86.3% [78.3, 91.6] | 82.4% [74.3, 85.8] |
|  | 3 – <6y | 76.9% [65.4, 85.5] | 70.5% [62.9, 75.5] |
|  | 6 – <18y | 71.3% [61.0, 79.7] | 68.1% [60.8, 72.9] |
|  | 18 – 39 | 28.4% [19.4, 39.5] | 43.8% [40.0, 46.2] |
|  | 40 – 59y | 8.2% [3.6, 17.8] | 9.6% [9.0, 10.0] |
| 5 Doses (Primary series + early childhood booster + childhood booster)* |  |  |  |
| Pachon et al. (2002) [29]<br>Setting: Spain<br>Ages included: 2–39 years<br>Cross-sectional study (n = 3,932) | 2 – 5y | 90% [85.0, 90.0] | 73.0% [64.0, 78.3] |
|  | 6 – 9y | 98.6% [96.6, 100] | 62.2% [54.6, 67.6] |
|  | 15 – 19y | 88% [83.0, 93.0] | 60.6% [53.6, 65.4] |
|  | 25 – 29y | 67% [62.0, 72.0] | 48.1% [43.2, 51.6] |
|  | 30 – 39y | 42% [38.0, 47.0] | 32.6% [29.4, 34.8] |

Notes: Values in brackets show the 95% confidence interval for the study values and the model run for the lower and upper range of duration of population protection. <sup>^</sup>This is an average across the three 5-year bands presented in the original study as historic vaccination coverage varied. In the simulation. \*Confidence intervals

For the primary series, the model estimates population protection that is 4.5% lower than the seroprevalence study [27]. However, the model output falls within the seroprevalence CI range and vice versa. For the four-dose setting (China), model and study values are very close for all but the 18–39 years of age category, where it is 4.3% higher than the upper seroprevalence estimate [28]. The values for the other age groups are all within the 95%CI bands and only differ to the central estimate by 3.7% on average. For the 5-dose setting (Spain), the model underestimates the level of protection across all age groups [29]. While this analysis ran the model using exponential waning, for settings where seroprevalence studies are available, the waning function can be fitted to better replicate the decline. Overall, our model reasonably replicates the immunity levels and pattern of decline across all three settings, although underestimates the immunity level compared to Pachon et al. [29].

## 4. Discussion

### 4.1. Key findings and features

We have developed an age-structured, deterministic compartmental model of tetanus. We conducted a sensitivity analysis and explored the immunity dynamics of the model. We found that the model is most sensitive to changes in the transmission parameter, likelihood of death, and the duration of protection from vaccination (immunity waning rate). The first two are unlikely to substantially impact model results as 1) the transmission scalar is a model derived parameter, which is determined during calibration, and 2) tetanus mortality rates are usually well estimated, owing to the severe nature of the disease.

The modelling of immune waning is a crucial component in understanding the dynamics of infectious diseases, particularly in the context of vaccination. Given the sensitivity of the immunity waning rate on key model outputs (especially incidence and population protection) across ages of interest, we validated the model’s projections of population protection using three seroprevalence studies [27–29]. Our model replicates the qualitative pattern of tetanus immunity shown in Figure 2.4 [2,5,13] and is able to produce population protection estimates that align with seroprevalence studies in settings with different vaccination schedules and coverage [27,28].

Different approaches to modelling immune waning can lead to varying implications for disease control strategies, depending on the nature of the disease and the population under consideration. We explored two approaches to modelling the decline of vaccine-acquired immunity and proceeded with an exponential waning rate. Exponential waning, one of the most common approaches, assumes a constant decay rate of immunity, making it computationally simple and suitable for long-term vaccine efficacy estimation [30,31]. However, this method overlooks individual variability in immune responses, limiting its ability to capture the heterogeneity of immunity loss across populations [32]. Other approaches, such as piecewise or stepwise waning, account for changes in decay rates at different stages, potentially offering a more realistic description of immune dynamics post-vaccination. While these models may be able to better reflect different observed patterns of immunity loss, their complexity requires more detailed data and assumptions about the timing of immunity transitions, which is limited in the literature. Ultimately, the choice of immune waning model is context-dependent, with trade-offs between realism, computational feasibility, and available data to inform the waning decay function, influencing the selection of an appropriate approach.

### 4.2. Contributions

There are very few tetanus models available in the literature. This model adds to the body of knowledge and makes several advancements. The most important contribution is the inclusion of distinct states for each vaccine dose, which is especially valuable in the context of exploring the impact of vaccination strategies, such as the introduction of different combinations of DTPCV booster doses. While this increases the number of compartments and model complexity, it allows the cumulative effectiveness and protection of each subsequent dose to be modelled explicitly, and for the model to maintain immune history.

This model also makes advancements on existing models by capturing detailed age breakdowns. This enables 1) precise vaccine schedules to be considered, and 2) declining immunity over age to be modelled. Where local seroprevalence studies are available, the immune profile by age can be fitted to this data. A future advancement in the model could be the inclusion of age-specific transmission rates, to capture high risk groups (e.g. adolescent boys undergoing male circumcision).

The model includes specific immune profiles, incidence and mortality for maternal and neo-natal tetanus. Neo-natal tetanus can be included in the model, depending on the local setting, such as the level of bacteria in the environment and whether MNTE has been achieved. There is also the option to model the influence of public health measures like safe birth practices for reducing NT tetanus.

### 4.3. Limitations

Although the model captures dose-specific levels of immunity, protection is modelled in a binary manner (i.e. protected or not-protected) and does not account for grades of immunity over time following vaccination. Although this is reasonable given the lack of conclusive literature on the nature of immune waning, an exploration of the structural uncertainty both in terms of the waning function (as discussed above) and potential grades of waned protection, would be an interesting avenue of future research.

While the detailed age groups and dose-specific protection in the model enables calibration to seroprevalence data, there are limitations to this approach. Firstly, it is not straightforward to map the modelled protection level to an equivalent seroprevalence cutoff. Secondly, the lack of standardised methods, differences in findings across settings even with the same schedule and similar coverage, and limited availability of high-quality representative serosurveys raise challenges for comparison and interpretation [33].

A common challenge encountered in the development and validation of mathematical models is variability and uncertainty in the available data. Due to the severe nature of the disease, the true burden of tetanus is better understood than for many other diseases. However, establishing the true burden is still prone to challenges. In epidemiological studies, the quality of data is often compromised by inconsistencies in reporting, incomplete case records, and differences in diagnostic criteria across regions and time periods [34]. Further, the scarcity of data for certain populations or settings limits the ability to fully capture the dynamics of tetanus transmission, especially in low-resource or remote areas. Fluctuations in vaccination coverage, healthcare access, and environmental factors are often difficult to quantify accurately, contributing to model uncertainty. While we have tried to address this by presenting results with uncertainty ranges, the model’s reliability is inherently constrained by the quality and completeness of the underlying data. This limitation is mitigated in the application of the model, as scenario analysis allows for the relative comparison of different strategies without the focus being on forecasting precise numbers of cases or protection.

## 5. Conclusion

We have developed a novel age-structured, deterministic compartmental model of tetanus transmission and vaccination. The model is driven by environmental transmission and incorporates interventions like hygiene and safe birth practices to reduce the magnitude of environmental transmission. It explicitly models vaccination, separating each dose of the primary series, booster doses, and maternal vaccination to capture dose-specific effectiveness and duration of protection. The model captures heterogeneous immunity profiles by dose and age, and the cumulative nature of vaccine-derived protection. The immune dynamics follow the patterns described in literature and can replicate seroprevalence studies, although the exact characterisation of immunity in the literature still has gaps. This model presents a substantial advancement on previously published models and is well positioned to inform tailored vaccination strategies to reduce neonatal and non-neonatal tetanus.

## Author contributions

Rachel A. Hounsell: Conceptualization; Data curation; Formal analysis; Funding acquisition; Investigation; Methodology; Project administration; Validation; Visualization; Writing - original draft. Jared Norman: Conceptualization; Formal analysis; Methodology; Writing - review & editing. Sheetal P. Silal: Conceptualization; Funding acquisition; Methodology; Supervision; Validation; Writing - review & editing.

## Data availability

All relevant data are publicly available or within the Appendices.

## Abbreviations and Acronyms

CFR: Case fatality rate
CI: Confidence interval
DTPCV: Diphtheria, tetanus, and pertussis-containing vaccine
EC: Early childhood booster (scenario abbreviation)
ELISA: Enzyme-linked immunosorbent assay
EPI: Expanded programme on immunisation
GBD: Global burden of disease
IgG: Immunoglobulin G
LHS: Latin hypercube sampling
LMIC: Low- and middle-income countries
MNT: Maternal and neonatal tetanus
MNTE: Maternal and neonatal tetanus elimination
NT: Neonatal tetanus
PRCC: Partial rank correlation coefficient
SDI: Sociodemographic index
SEIRD: Susceptible-exposed-infected-recovered-deceased
TEG: Technical expert group
TT: Tetanus toxoid
TTCV: Tetanus-toxoid-containing vaccines
UNFPA: United Nations Population Fund
WHO: World Health Organisation

## Acknowledgements

We would like to extend our thanks to the Technical Expert Group who served as scientific advisors and provided input on model development, epidemiology and immunology of disease: Dr Ampeire Immaculate, Ugandan National Immunization Program, Ministry of Health, Government of Uganda; Dr Anna Acosta, Meningitis and Vaccine Preventable Diseases Branch at U.S. Centers for Disease Control and Prevention (CDC); Dr Annet Kisakye, Expanded Program on Immunization, World Health Organization, Uganda; Prof Paula Mendes Luz, Instituto Nacional de Infectologia Evandro Chagas (INI); Dr Todi Mengistu, Measurement, Evaluation and Learning (MEL) team at Gavi, the Vaccine Alliance; Dr Helen Quinn, National Centre for Immunisation Research and Surveillance (NCIRS); University of Sydney Children’s Hospital Westmead Clinical School; So Yoon Sim, Value of Vaccines, Modeling & Economics (VoV) team, Immunization Analysis & Insights (IAI) unit, Department of Immunization, Vaccines and Biologicals (IVB), World Health Organization; Dr Rania Tohme, Hepatitis B and Tetanus Team in the Global Immunization Division at the U.S. Centers for Disease Control and Prevention (CDC); and Prof Rudzani Muloiwa, Vaccines for Africa Initiative (VACFA), Faculty of Health Sciences, University of Cape Town, South Africa. We would also like to extend our thanks to Mieke du Plessis, for her valuable support proofreading the manuscript.

## Appendices – Tetanus Model Details

### Age groups

**Table A.1.**
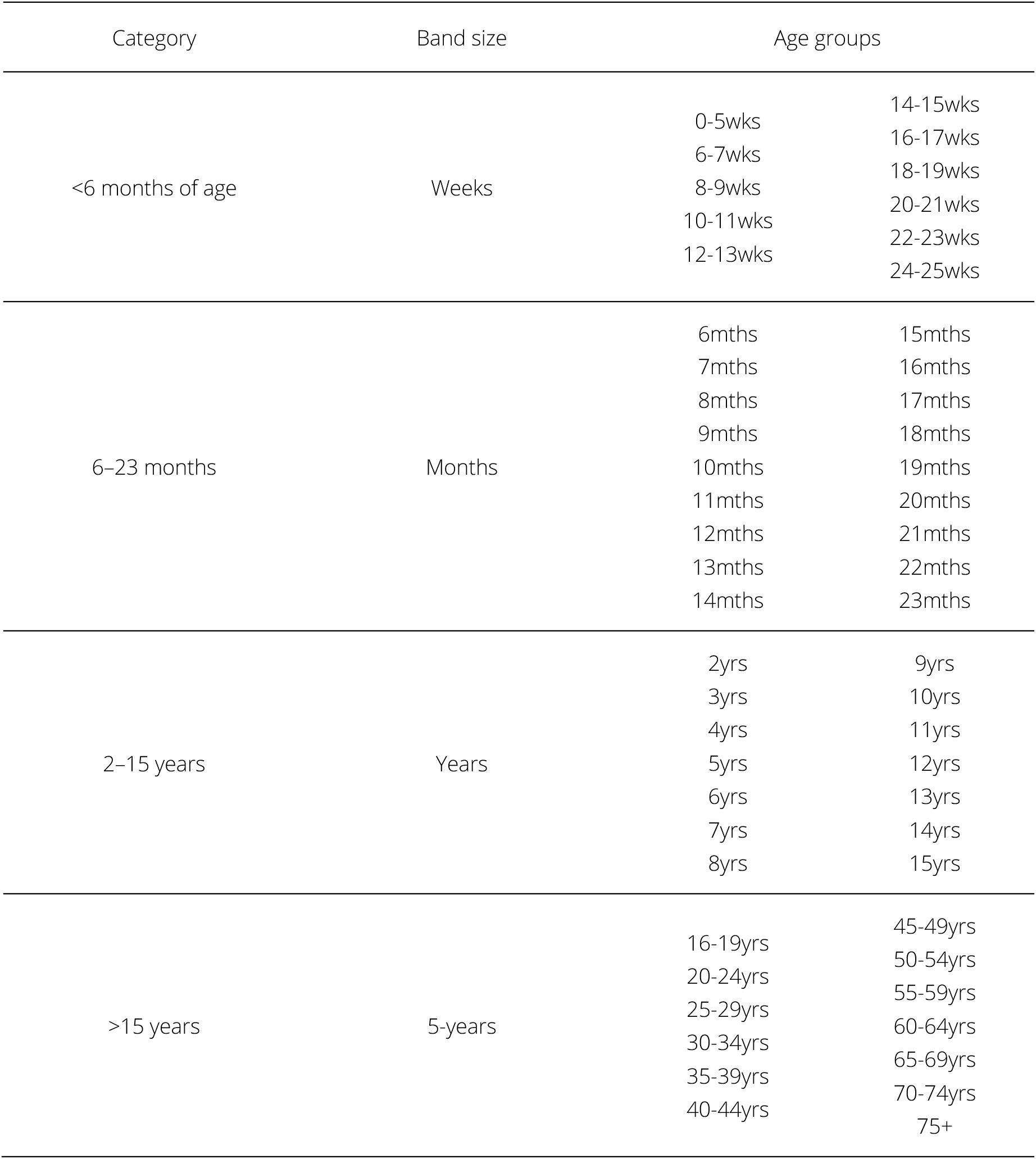
Model age groups.

### Model structure

**Table A.2.**
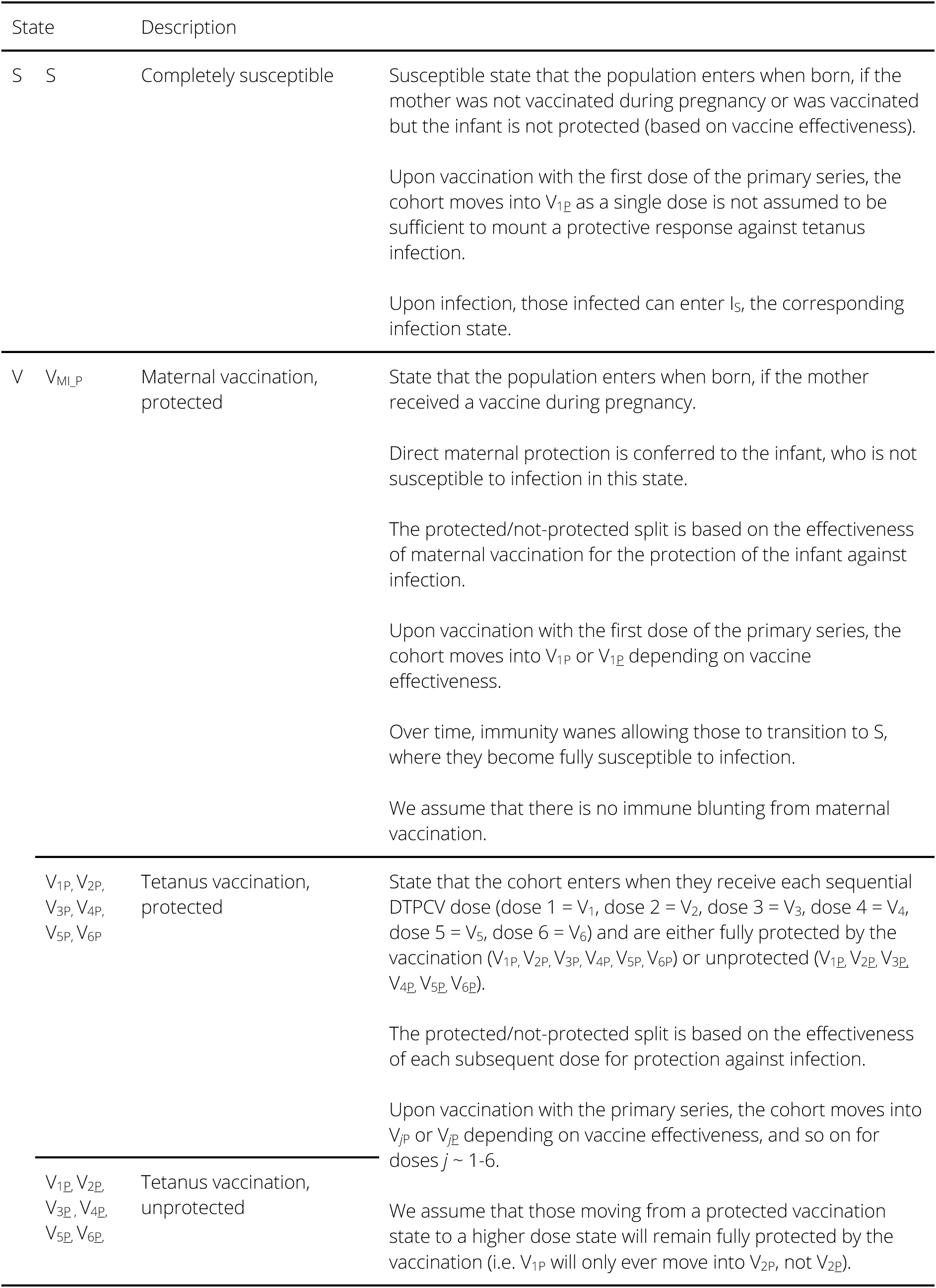
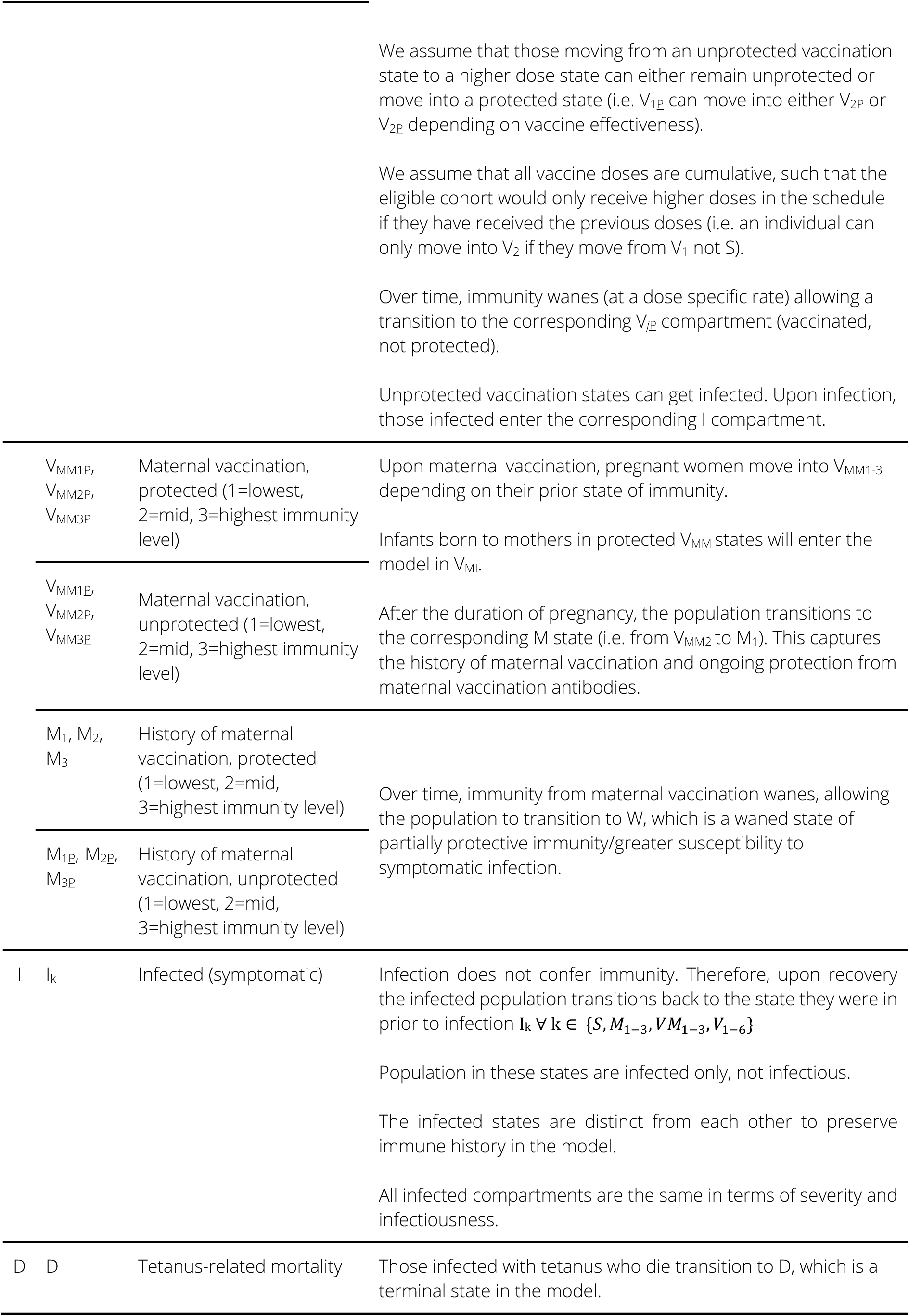
Tetanus model compartments (states), descriptions and key assumptions.

### Model equations

The rates of change of the population in each epidemiological state in the tetanus model are described by the following set of ordinary differential equations, for each age group *i* = 56:

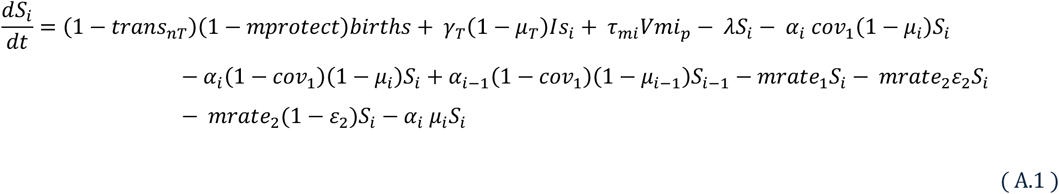

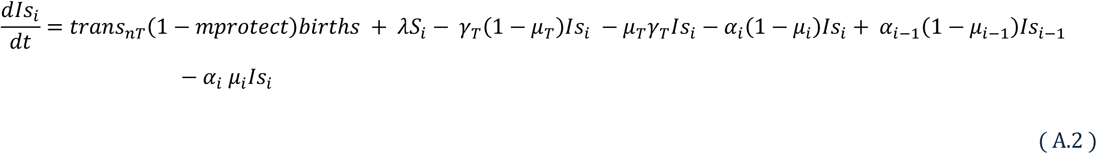

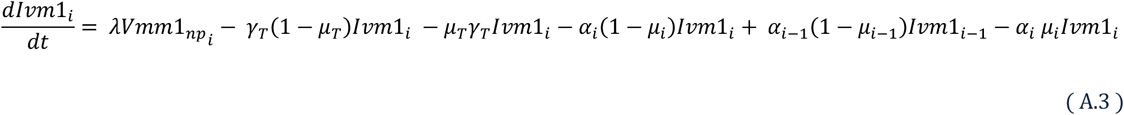

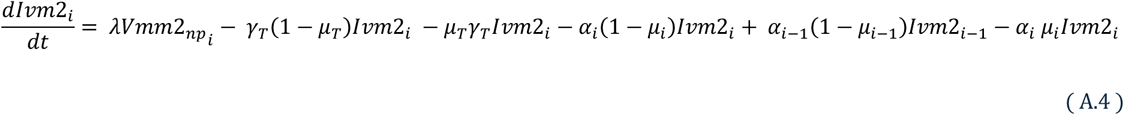

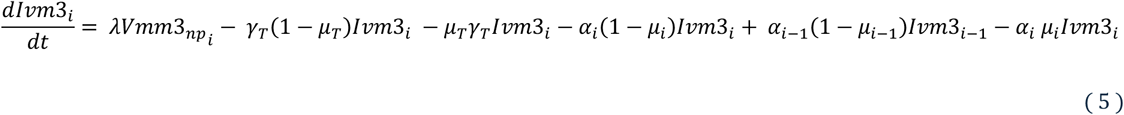

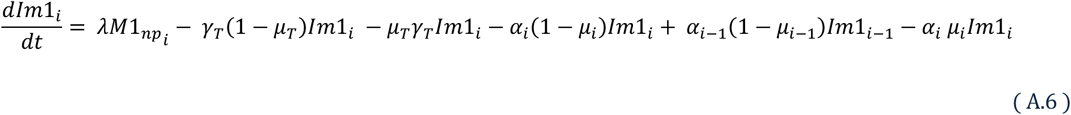

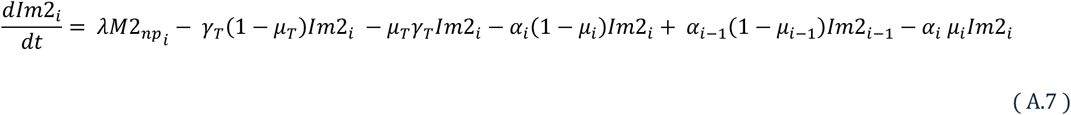

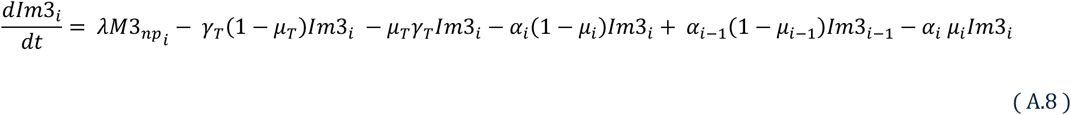

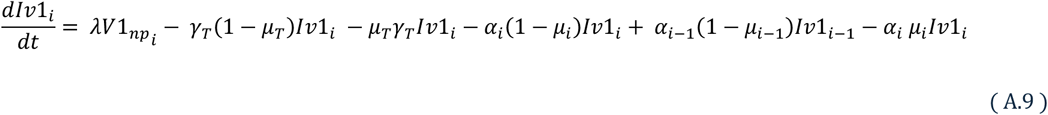

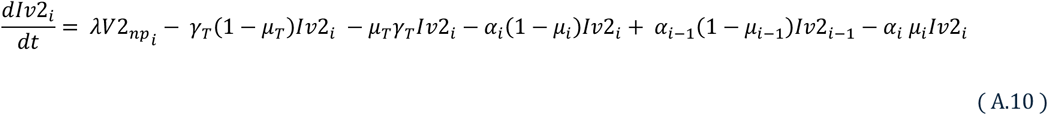

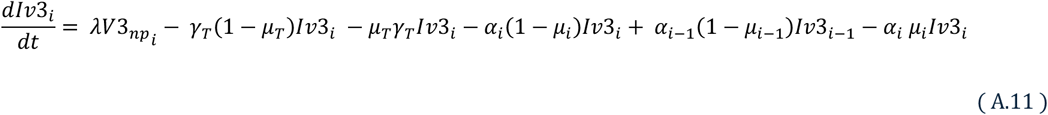

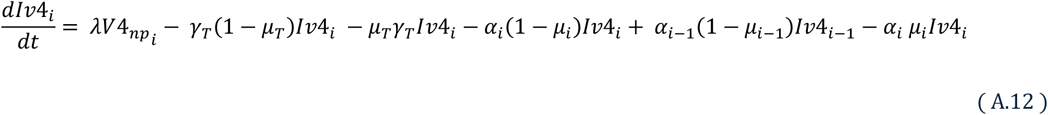

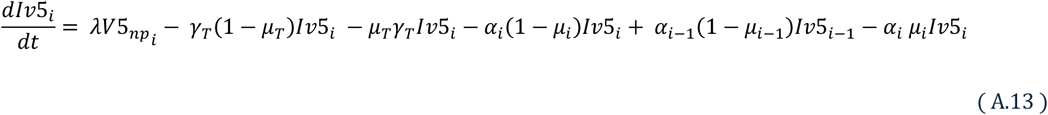

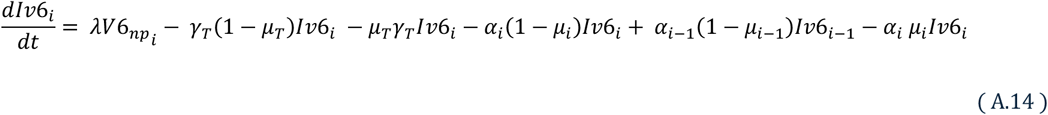

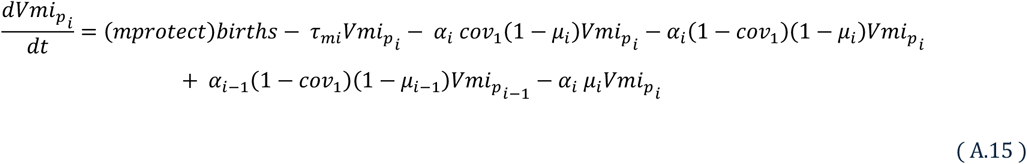

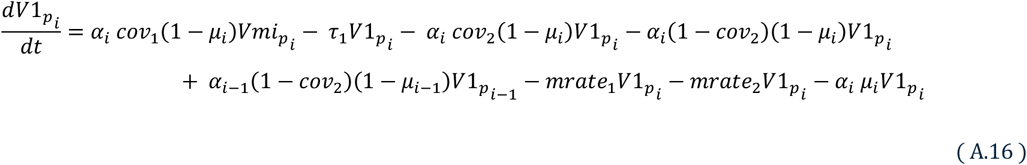

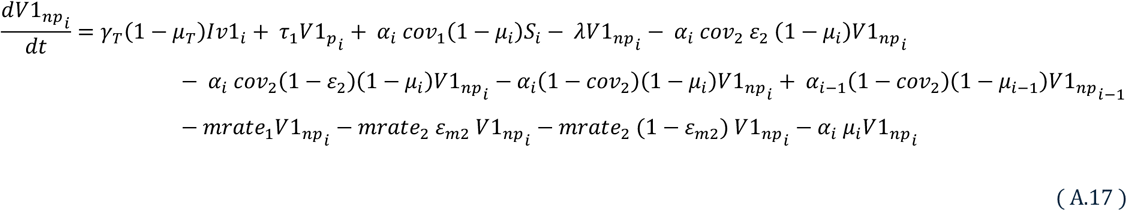

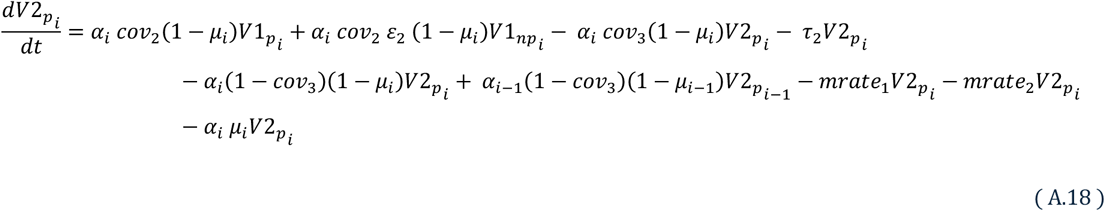

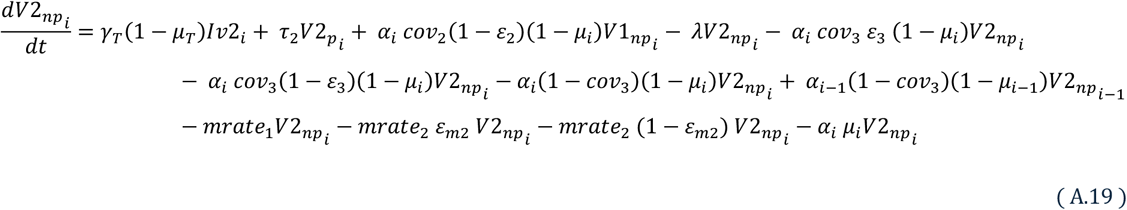

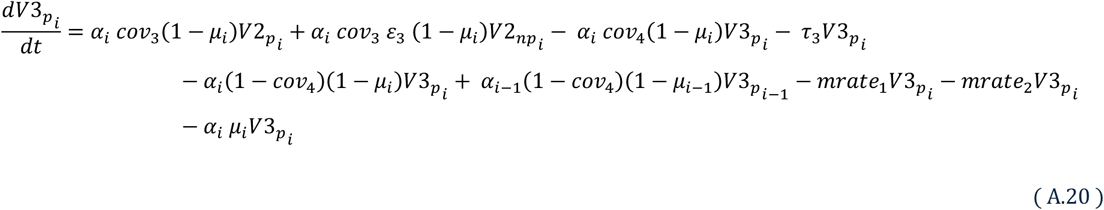

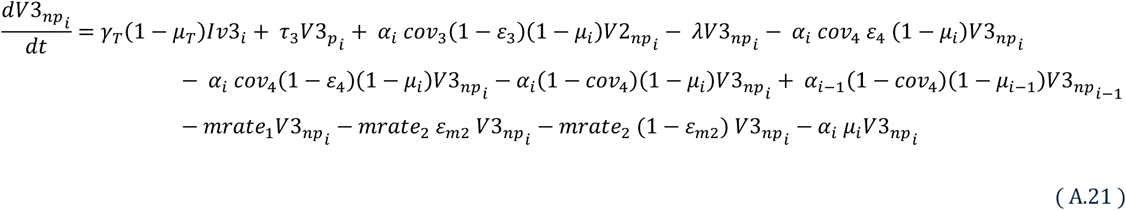

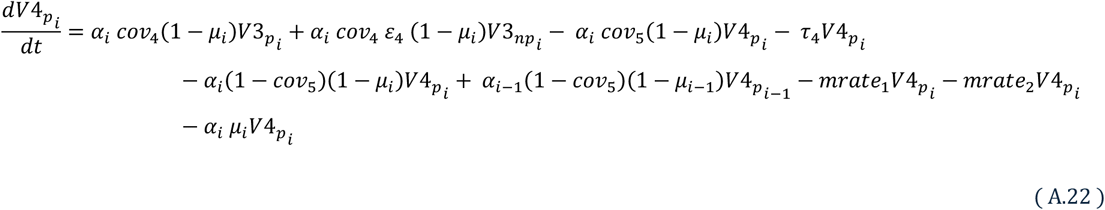

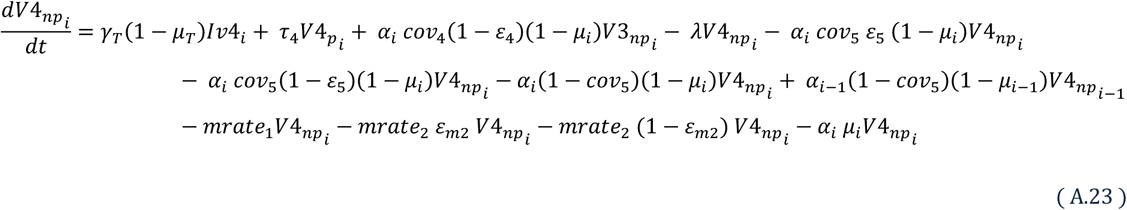

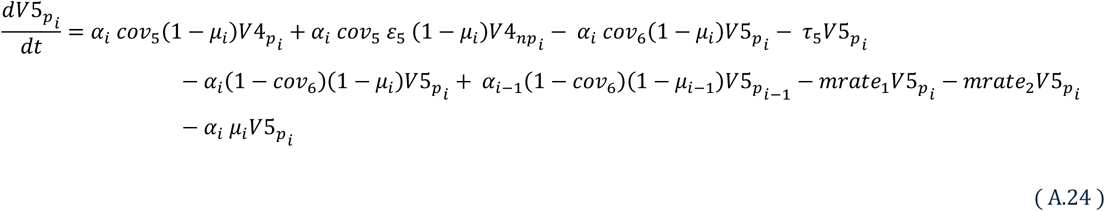

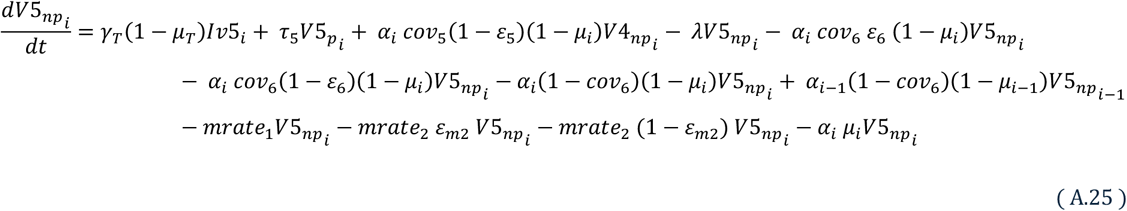

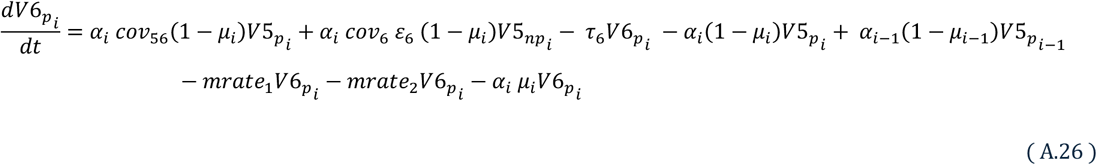

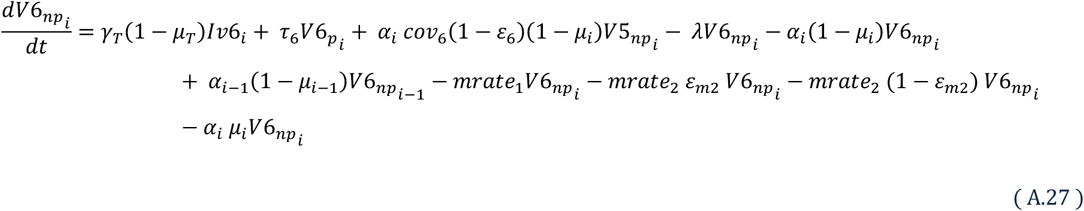

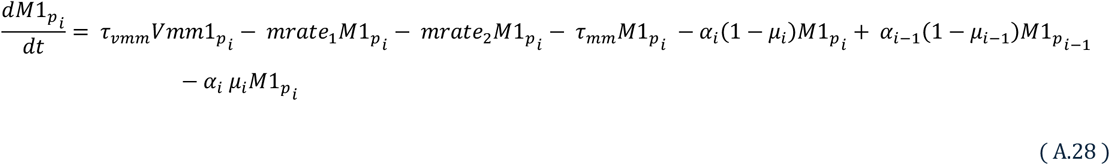

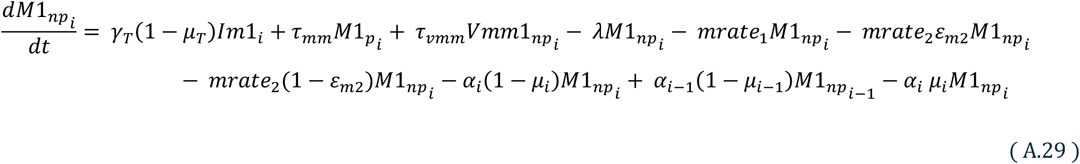

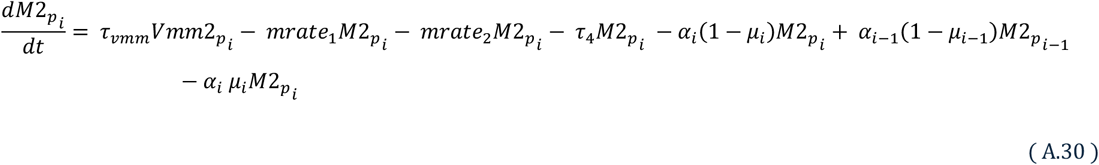

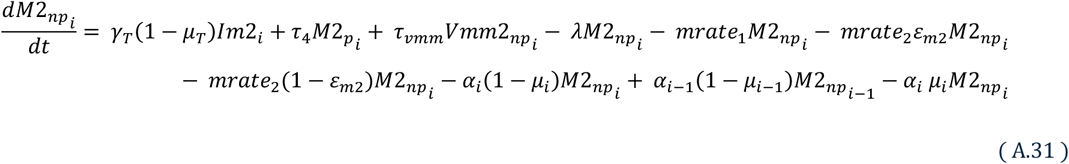

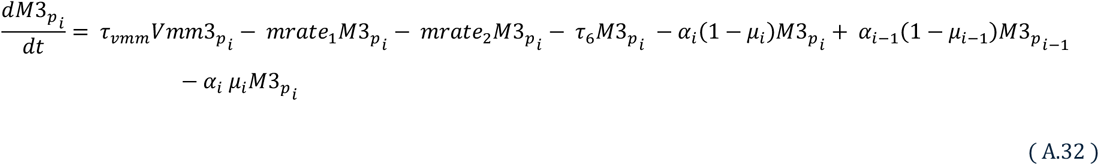

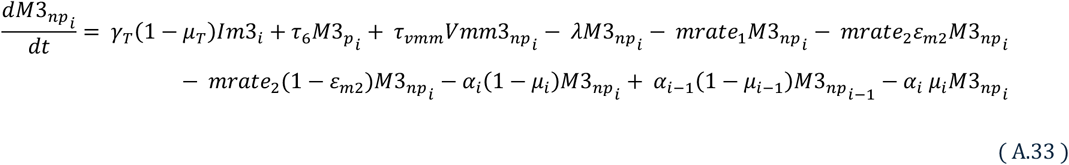

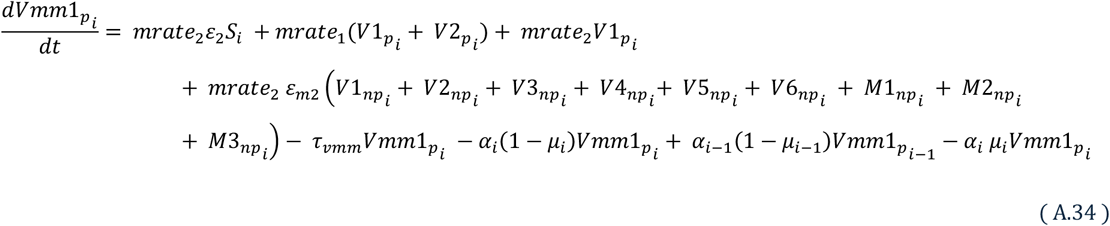

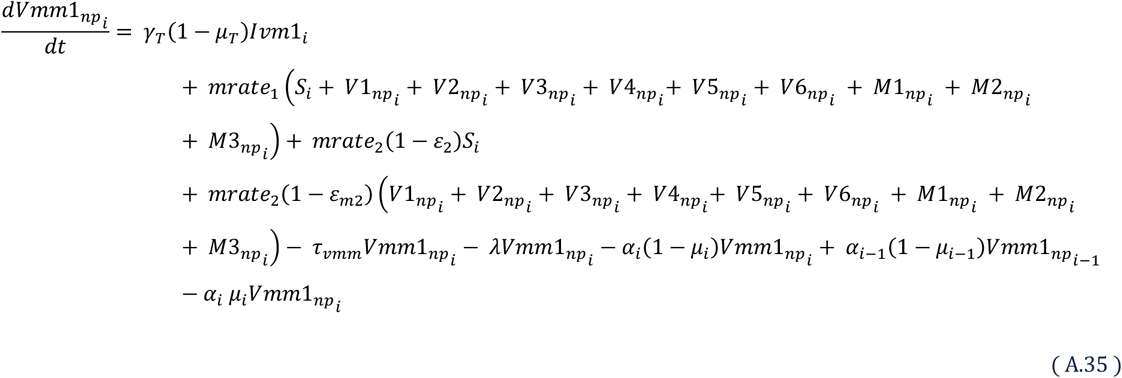

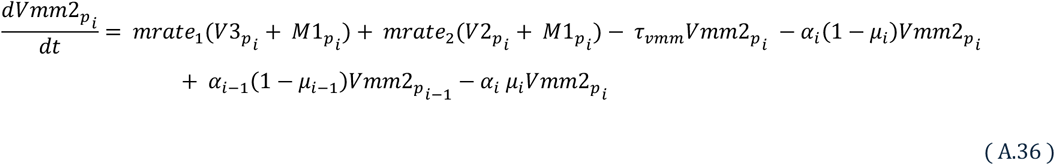

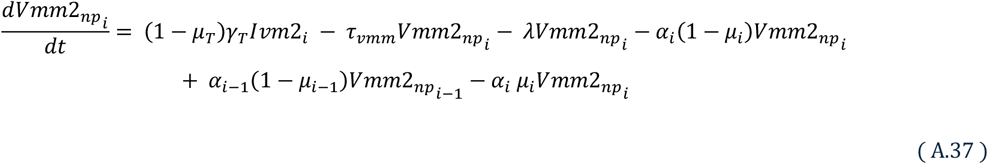

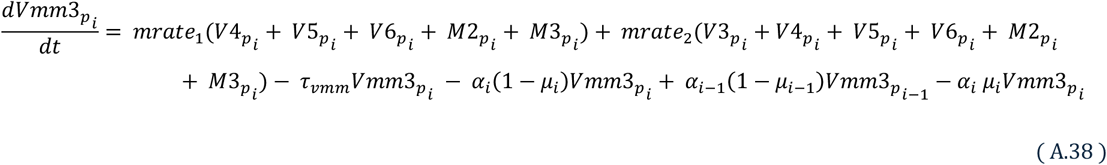

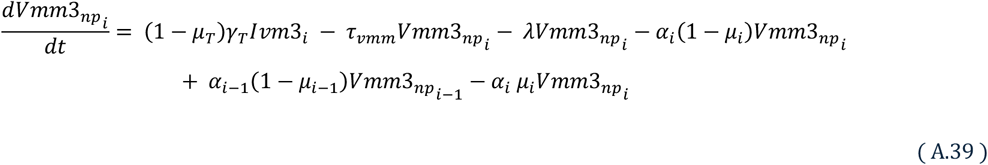

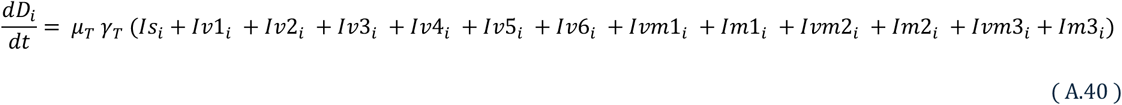

### Parameters

**Table A.3.**
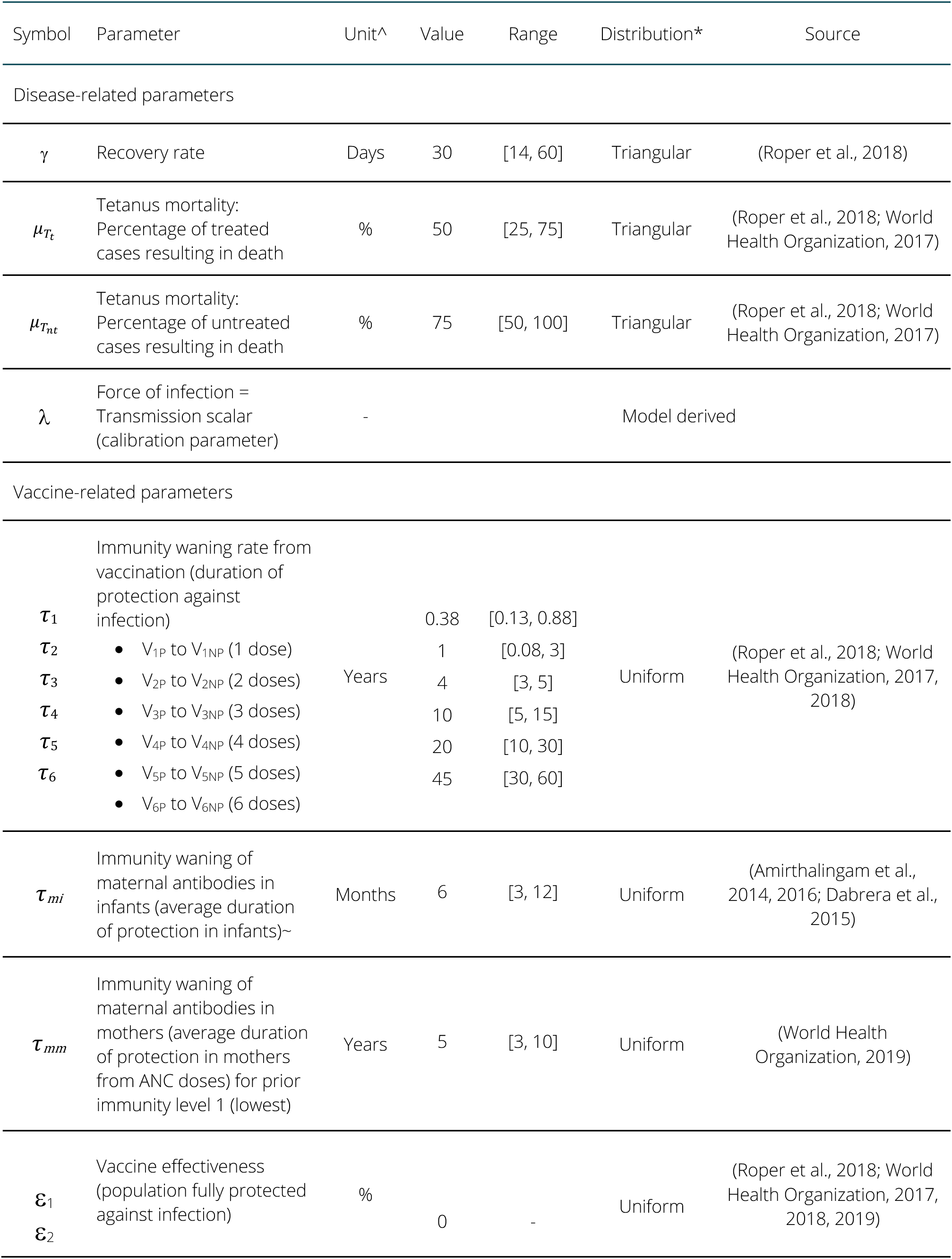
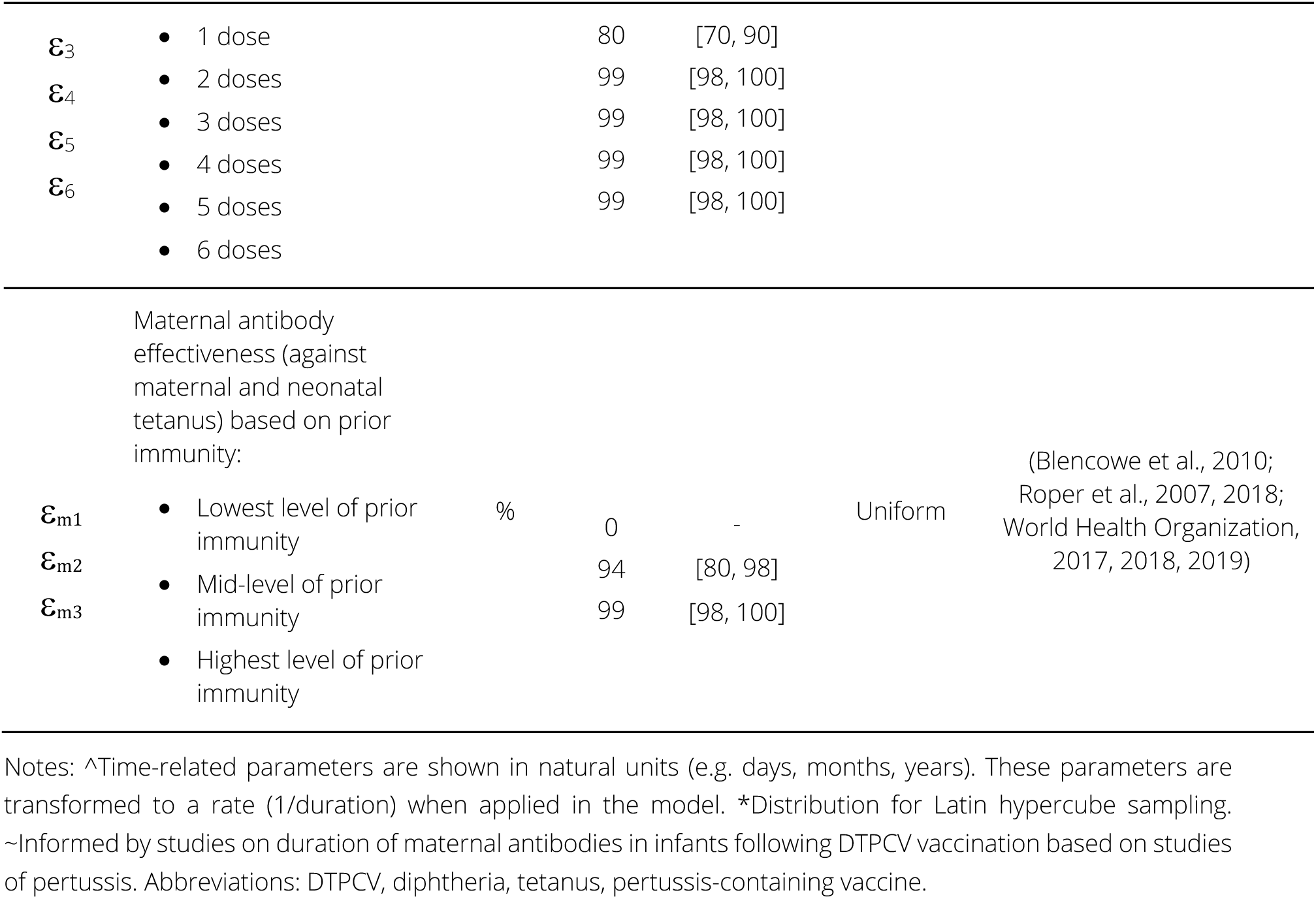
Model parameters for tetanus.

### Technical Expert Group

The role of the technical expert group (TEG) was to advise on the model development, epidemiology and immunology of disease, and broader project guidance. The parameter values were established with input and review from the TEG.

- **Dr Ampeire Immaculate**, Ugandan National Immunization Program, Ministry of Health, Government of Uganda.
- **Dr Anna Acosta**, Meningitis and Vaccine Preventable Diseases Branch at U.S. Centers for Disease Control and Prevention (CDC)
- **Dr Annet Kisakye**, Expanded Program on Immunization, World Health Organization, Uganda
- **Prof Paula Mendes Luz**, Instituto Nacional de Infectologia Evandro Chagas (INI)
- **Dr Todi Mengistu**, Measurement, Evaluation and Learning (MEL) team at Gavi, the Vaccine Alliance
- **Dr Helen Quinn**, National Centre for Immunisation Research and Surveillance (NCIRS); University of Sydney Children’s Hospital Westmead Clinical School.
- **So Yoon Sim**, Value of Vaccines, Modeling & Economics (VoV) team, Immunization Analysis & Insights (IAI) unit, Department of Immunization, Vaccines and Biologicals (IVB), World Health Organization
- **Dr Rania Tohme**, Hepatitis B and Tetanus Team in the Global Immunization Division at the U.S. Centers for Disease Control and Prevention (CDC)

